# Association Between Preoperative Anemia and Postoperative Outcomes in Adults Undergoing Cardiac Surgery: A Systematic Review and Meta-analysis

**DOI:** 10.64898/2026.09.24.26363864

**Authors:** Wesam Abedalqader Hammad, Abdalrahman Hammad

**Affiliations:** Faculty of Medicine, Sumy State University, Sumy, Ukraine; Faculty of Medicine, Zagazig University, Zagazig, Egypt

**Keywords:** cardiac surgery, preoperative anaemia, acute kidney injury, red-cell transfusion, perioperative outcomes

## Abstract

Preoperative anaemia is common in adults undergoing cardiac surgery and may identify patients at increased perioperative risk. This systematic review and meta-analysis synthesised contemporary observational evidence in accordance with PRISMA 2020. Five databases were searched for peer-reviewed English language studies published from 2017 to July 2026. Random effects meta-analyses used restricted maximum likelihood estimation with Hartung-Knapp confidence intervals; crude exact count estimates were synthesised primarily and adjusted estimates separately. Fourteen observational cohort studies involving 74,571 adults, including 17,349 (23.3%) with anaemia, were included. Preoperative anaemia was associated with red cell transfusion (OR, 3.66; 95% CI, 2.14-6.24), acute kidney injury or renal failure (OR, 2.43; 95% CI, 1.84-3.21), early mortality (OR, 2.11; 95% CI, 1.40-3.17), and postoperative infection (OR, 1.49; 95% CI, 1.21-1.82), but not postoperative stroke (OR, 1.16; 95% CI, 0.43-3.17). Certainty was low for renal outcomes and very low for the other outcomes. Preoperative anaemia is an important perioperative risk marker, particularly for transfusion and renal complications, although causality cannot be established.

## 1. INTRODUCTION

Cardiovascular diseases remain the leading global cause of death and a major contributor to morbidity (World Health Organization, 2025). Cardiac surgery remains an important treatment for selected patients with advanced coronary and valvular disease, including surgical coronary revascularisation and valve repair or replacement (Vrints et al., 2024; Praz et al., 2025). Although operative technique, anaesthesia, intensive care, and patient blood-management programmes have improved, perioperative anaemia and red-cell transfusion remain frequent and clinically consequential features of cardiac surgery (Howard-Quijano et al., 2026).

Preoperative anaemia is common among patients undergoing cardiac surgery, but reported prevalence varies substantially with age, comorbidity burden, procedure type, recruitment setting, and the threshold used to define anaemia (Howard-Quijano et al., 2026; Frei et al., 2025; Zhang et al., 2026). The 2024 World Health Organization guideline defines anaemia in adults as haemoglobin below 13.0 g/dL in men and below 12.0 g/dL in non-pregnant women (World Health Organization, 2024). Cardiac-surgical cohorts, however, also use haematocrit thresholds, iron-status criteria, or modified definitions, creating genuine clinical and statistical heterogeneity across studies (Alkadri et al., 2026; Zhang et al., 2026).

The association between anaemia and postoperative outcomes has several plausible biological pathways. Reduced haemoglobin lowers arterial oxygen content and may impair tissue oxygen delivery during cardiopulmonary bypass, haemodilution, hypotension, and other periods of perioperative stress (Chin et al., 2023). Anaemia also increases the probability of red-cell transfusion, while transfusion itself may be associated with immunomodulation, pulmonary complications, and other adverse outcomes (Howard-Quijano et al., 2026). These pathways are not independent of baseline risk: anaemia frequently coexists with older age, chronic kidney disease, diabetes, heart failure, and other comorbidities that independently predict acute kidney injury, infection, stroke, and mortality (Ripoll et al., 2021). Consequently, an observed association between anaemia and outcome can reflect both a biological contribution of reduced oxygen-carrying capacity and confounding by the clinical conditions that produced or accompany anaemia.

The distinction between association and causation is particularly important in observational cardiac-surgery evidence. A crude comparison may capture the total difference in postoperative events between anaemic and non-anaemic patients, but it does not determine whether correcting haemoglobin would eliminate that difference. Published adjusted estimates can address some confounding, yet the covariates, definitions, and modelling strategies are rarely identical across cohorts. A rigorous synthesis therefore needs to report crude and adjusted evidence separately, examine heterogeneity rather than conceal it, and downgrade certainty when residual confounding or imprecision remains.

In an earlier systematic review and meta-analysis of 22 observational studies involving 114,277 patients, Padmanabhan et al. reported associations between preoperative anaemia and mortality, acute kidney injury, stroke, postoperative infection, and red-cell transfusion (Padmanabhan et al., 2019). That review searched the literature through 2017. Since then, additional cohorts have examined transfusion, renal outcomes, mortality, stroke, and infection in contemporary cardiac-surgery populations (Alkadri et al., 2026; Shales et al., 2024). These newer studies differ in procedure mix, anaemia definition, endpoint timing, use of propensity matching, and adjustment for comorbidity, making an updated synthesis necessary.

The present review was designed to address that gap. The primary outcome was early mortality, defined as death during the index hospitalisation or within 30 days of surgery. Secondary outcomes were acute kidney injury or renal failure, perioperative red-cell transfusion, postoperative infection, and stroke. The review also prespecified sensitivity analyses for propensity-matched cohorts, study quality, exact event counts, and leave-one-out influence, together with a GRADE assessment that treats residual confounding as a property of the evidence rather than assuming that a high summary quality score removes it.

Accordingly, this systematic review and meta-analysis evaluated the magnitude, consistency, and certainty of associations between preoperative anaemia and postoperative outcomes in adults undergoing cardiac surgery, including studies published from 2017 through July 2026. The results are reported in accordance with PRISMA 2020 (Page et al., 2021).

## 2. METHODS

### 2.1 Study Design and Reporting Framework

This systematic review and meta-analysis evaluated the association between preoperative anemia and postoperative outcomes in adults undergoing cardiac surgery. The study was reported in accordance with the **Preferred Reporting Items for Systematic Reviews and Meta-Analyses (PRISMA) 2020 statement (Page et al., 2021).** The review protocol was registered in PROSPERO (registration number: CRD420261472633).

### 2.2 Search Strategy

PubMed/MEDLINE, Scopus, Embase, Web of Science, and the Cochrane Library were searched on 20 July 2026 for English-language studies published from 1 January 2017 through 20 July 2026, using database-specific strategies combining terms related to preoperative anaemia, cardiac surgery, and postoperative outcomes. The searches incorporated relevant alternative spellings where applicable. A supplementary verification search of journal websites was conducted on 18 August 2026, through which one additional eligible report was identified. This supplementary search did not alter the predefined primary database search period. Full database-specific search strategies are provided in the Supplementary Material (eTables S2a–e).

### 2.3 Eligibility Criteria

Studies were eligible if they included adults aged ≥18 years undergoing cardiac surgery; assessed preoperative anaemia before surgery; included a non-anaemic comparison group; and reported at least one relevant postoperative outcome. Eligible procedures included coronary artery bypass grafting (CABG), valve surgery, combined CABG and valve surgery, redo cardiac surgery, and adult congenital cardiac surgery. Outcomes included mortality, acute kidney injury, stroke, red-cell transfusion, postoperative infection, intensive care unit or hospital length of stay, and other major postoperative complications. Prospective or retrospective cohort studies and case-control studies published as full-text, peer-reviewed articles in English from 2017 through July 2026 were eligible. Studies were excluded if they involved paediatric or non-cardiac surgical populations; did not assess preoperative anaemia as the primary exposure; lacked a non-anaemic comparison group, relevant postoperative outcomes, or sufficient extractable data; or were case reports, case series, conference abstracts, editorials, letters, narrative reviews, systematic reviews, or meta-analyses. For duplicate reports from the same cohort, the most complete or relevant report was retained.

### 2.4 Study Selection Process

Study selection was performed independently by two reviewers. Duplicate records were identified and removed before screening. The remaining records were screened by title and abstract, followed by full-text assessment of potentially eligible studies against the predefined eligibility criteria. Rayyan, a web-based systematic review management tool, was used to facilitate the screening and study selection process (Ouzzani et al., 2016). Any disagreements between the reviewers were resolved through discussion and consensus.

### 2.5 Data Extraction and Management

Data were extracted using a predefined data-extraction form and recorded in structured electronic files using Microsoft Excel. One reviewer performed the initial data extraction, and a second reviewer verified the extracted data. Any discrepancies were resolved through discussion and consensus. The main characteristics of the included studies and the outcomes reported by each study are presented in Table 1.

**Table 1.**
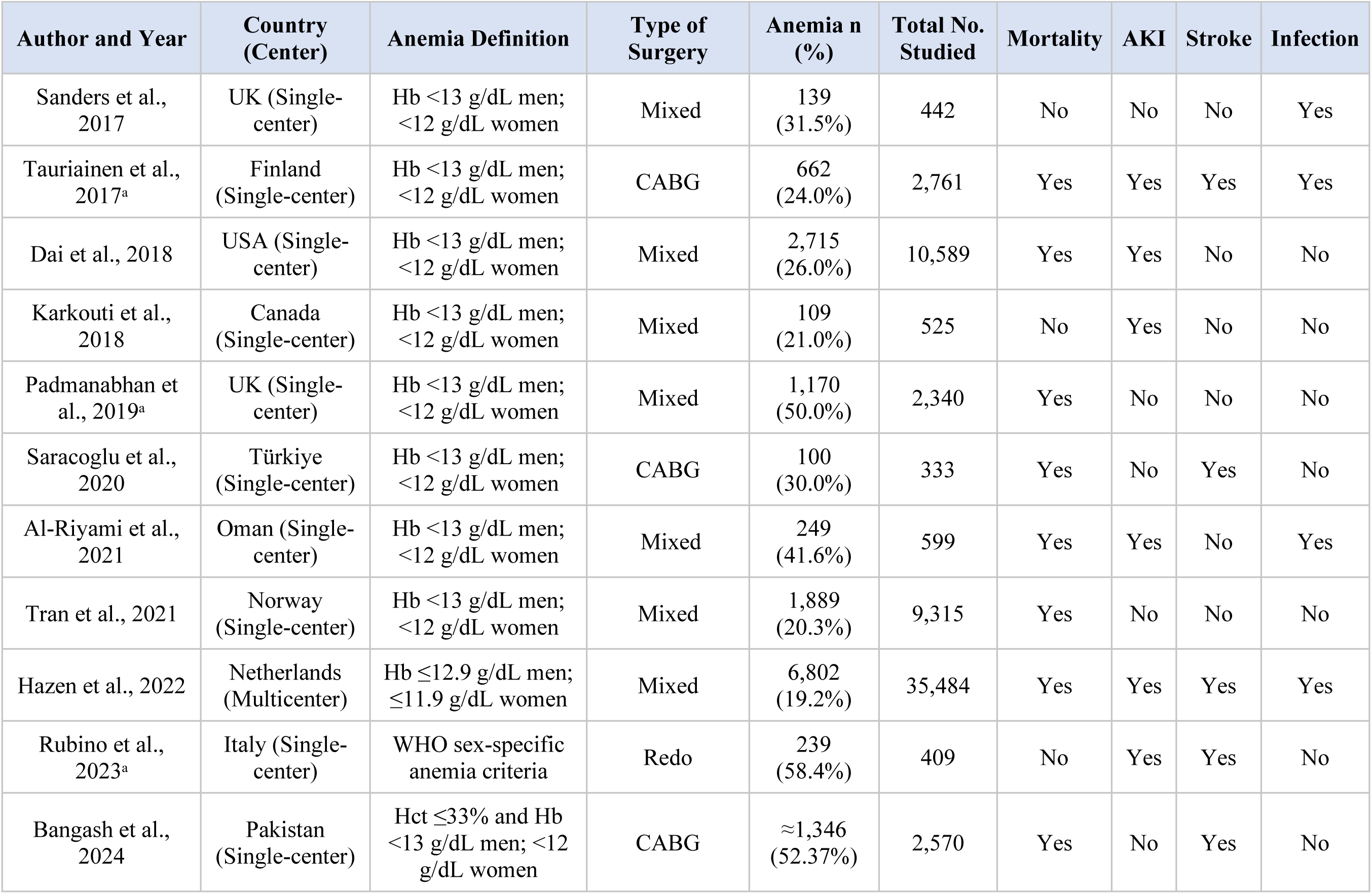
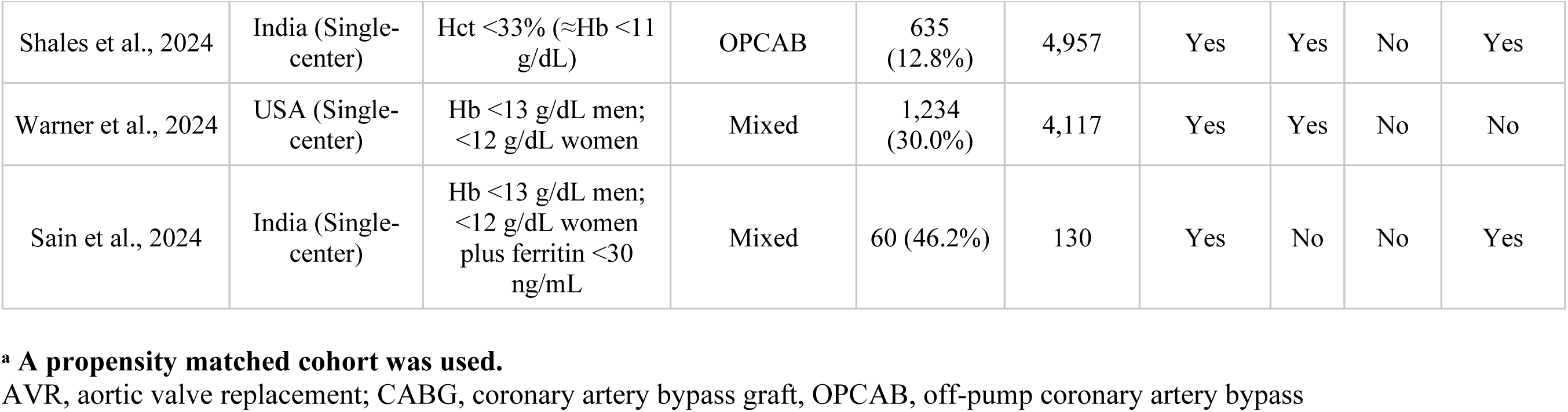
Characteristics of Included Studies.

| Author and Year | Country (Center) | Anemia Definition | Type of Surgery | Anemia n (%) | Total No. Studied | Mortality | AKI | Stroke | Infection |
| --- | --- | --- | --- | --- | --- | --- | --- | --- | --- |
| Sanders et al., 2017 | UK (Single-center) | Hb <13 g/dL men; <12 g/dL women | Mixed | 139 (31.5%) | 442 | No | No | No | Yes |
| Tauriainen et al., 2017<sup>a</sup> | Finland (Single-center) | Hb <13 g/dL men; <12 g/dL women | CABG | 662 (24.0%) | 2,761 | Yes | Yes | Yes | Yes |
| Dai et al., 2018 | USA (Single-center) | Hb <13 g/dL men; <12 g/dL women | Mixed | 2,715 (26.0%) | 10,589 | Yes | Yes | No | No |
| Karkouti et al., 2018 | Canada (Single-center) | Hb <13 g/dL men; <12 g/dL women | Mixed | 109 (21.0%) | 525 | No | Yes | No | No |
| Padmanabhan et al., 2019<sup>a</sup> | UK (Single-center) | Hb <13 g/dL men; <12 g/dL women | Mixed | 1,170 (50.0%) | 2,340 | Yes | No | No | No |
| Saracoglu et al., 2020 | Türkiye (Single-center) | Hb <13 g/dL men; <12 g/dL women | CABG | 100 (30.0%) | 333 | Yes | No | Yes | No |
| Al-Riyami et al., 2021 | Oman (Single-center) | Hb <13 g/dL men; <12 g/dL women | Mixed | 249 (41.6%) | 599 | Yes | Yes | No | Yes |
| Tran et al., 2021 | Norway (Single-center) | Hb <13 g/dL men; <12 g/dL women | Mixed | 1,889 (20.3%) | 9,315 | Yes | No | No | No |
| Hazen et al., 2022 | Netherlands (Multicenter) | Hb ≤12.9 g/dL men; ≤11.9 g/dL women | Mixed | 6,802 (19.2%) | 35,484 | Yes | Yes | Yes | Yes |
| Rubino et al., 2023<sup>a</sup> | Italy (Single-center) | WHO sex-specific anemia criteria | Redo | 239 (58.4%) | 409 | No | Yes | Yes | No |
| Bangash et al., 2024 | Pakistan (Single-center) | Hct ≤33% and Hb <13 g/dL men; <12 g/dL women | CABG | ≈1,346 (52.37%) | 2,570 | Yes | No | Yes | No |
| Shales et al., 2024 | India (Single-center) | Hct <33% (≈Hb <11 g/dL) | OPCAB | 635 (12.8%) | 4,957 | Yes | Yes | No | Yes |
| Warner et al., 2024 | USA (Single-center) | Hb <13 g/dL men; <12 g/dL women | Mixed | 1,234 (30.0%) | 4,117 | Yes | Yes | No | No |
| Sain et al., 2024 | India (Single-center) | Hb <13 g/dL men; <12 g/dL women plus ferritin <30 ng/mL | Mixed | 60 (46.2%) | 130 | Yes | No | No | Yes |

<sup>a</sup> **A propensity matched cohort was used.**
AVR, aortic valve replacement; CABG, coronary artery bypass graft, OPCAB, off-pump coronary artery bypass

For dichotomous outcomes, event counts and total sample sizes were extracted separately for anaemic and non-anaemic groups. Where available, published covariate-adjusted odds ratios (ORs) and corresponding 95% confidence intervals (CIs) were also extracted.

### 2.6 Risk of Bias and Certainty of the Evidence

The methodological quality of the included observational studies was assessed independently by two reviewers using the Newcastle–Ottawa Scale (NOS) for cohort studies (Wells et al., n.d.; Stang, 2010). Studies were classified as having good (≥7), fair (5–6), or poor (<5) methodological quality according to their NOS scores. Disagreements were resolved by re-examination of the original study reports followed by discussion and consensus. Inter-reviewer agreement was assessed using percentage agreement and Cohen’s kappa (κ). Detailed Newcastle–Ottawa Scale assessments are provided in Supplementary eTable S5

Certainty of the evidence for the main outcomes was assessed using the Grading of Recommendations Assessment, Development and Evaluation (GRADE) approach (Guyatt et al., 2008). Detailed GRADE assessments are provided in Supplementary eTable S6.

### 2.7 Outcome Measures

The primary outcome of this systematic review was short-term mortality, defined as death within 30 days after cardiac surgery or during the index hospitalization.

Secondary outcomes included acute kidney injury or renal failure, postoperative stroke, perioperative or postoperative red blood cell transfusion, postoperative infection, intensive care unit (ICU) length of stay, hospital length of stay, and other major postoperative complications reported by eligible studies.

Where multiple definitions or time points were reported for an outcome, the definitions and time points used by each study were recorded and considered during data synthesis.

### 2.8 Statistical Analysis and Meta-analysis

Outcome data were analysed using random-effects meta-analysis to compare anaemic and non-anaemic patients. Dichotomous outcomes were summarised as odds ratios (ORs) with 95% confidence intervals (CIs). Between-study variance was estimated using restricted maximum likelihood (REML), and confidence intervals were calculated using the truncated Hartung–Knapp adjustment. Mantel–Haenszel and Peto common-effect estimates were also calculated as cross-checks. For studies with a single zero-event cell, a Haldane–Anscombe correction of 0.5 was applied, whereas studies with no events in either group were excluded from the pooled OR calculation.

The primary meta-analysis was prespecified as the crude exact-count synthesis because event counts were directly extractable and represented the most consistently comparable data available across studies. Covariate-adjusted estimates were synthesised separately because the adjustment sets, matching procedures, and modelling approaches differed between studies and could not be harmonised reliably. Matched cohorts were retained in the primary analysis, and sensitivity analyses excluding these cohorts were performed to assess the robustness of the findings. Heterogeneity was assessed using I², τ², and Cochran’s Q statistic (Higgins et al., 2003). I² values of <30%, 30–60%, and >60% were interpreted as low, moderate, and substantial heterogeneity, respectively. Ninety-five percent prediction intervals were calculated when at least five studies contributed to an outcome. Prespecified subgroup analyses were conducted according to surgery type, anaemia definition, outcome definition or timing, study design, geographic region, and cohort type. Sensitivity analyses included leave-one-out analysis, influence diagnostics, exclusion of propensity-matched cohorts, restriction to studies with Newcastle–Ottawa Scale scores of 8–9, and restriction to studies reporting exact event counts. For early mortality, additional sensitivity analyses were performed according to mortality timing, including in-hospital and 30-day mortality. Funnel plots and Egger’s test were not performed because no outcome included at least ten study estimates. Analyses were performed in R using the metafor package (Viechtbauer, 2010) and independently replicated in Stata; forest plots were generated for all pooled outcomes.

## 3. RESULTS

### 3.1 Study Selection

The primary database search conducted on 20 July 2026 yielded 1,316 records from the electronic databases. After 314 duplicates were removed, 1,002 titles and abstracts were screened. A total of 943 records were excluded as they were not relevant to the review, leaving 59 reports for full-text assessment. One additional report was identified through the supplementary verification search conducted on 18 August 2026 and was also assessed for eligibility, resulting in a total of 60 reports assessed at full text. After full-text assessment, 46 reports were excluded because they did not meet the eligibility criteria (Figure 1). Ultimately, 14 studies were included in the systematic review and meta-analysis.

**Figure 1.**
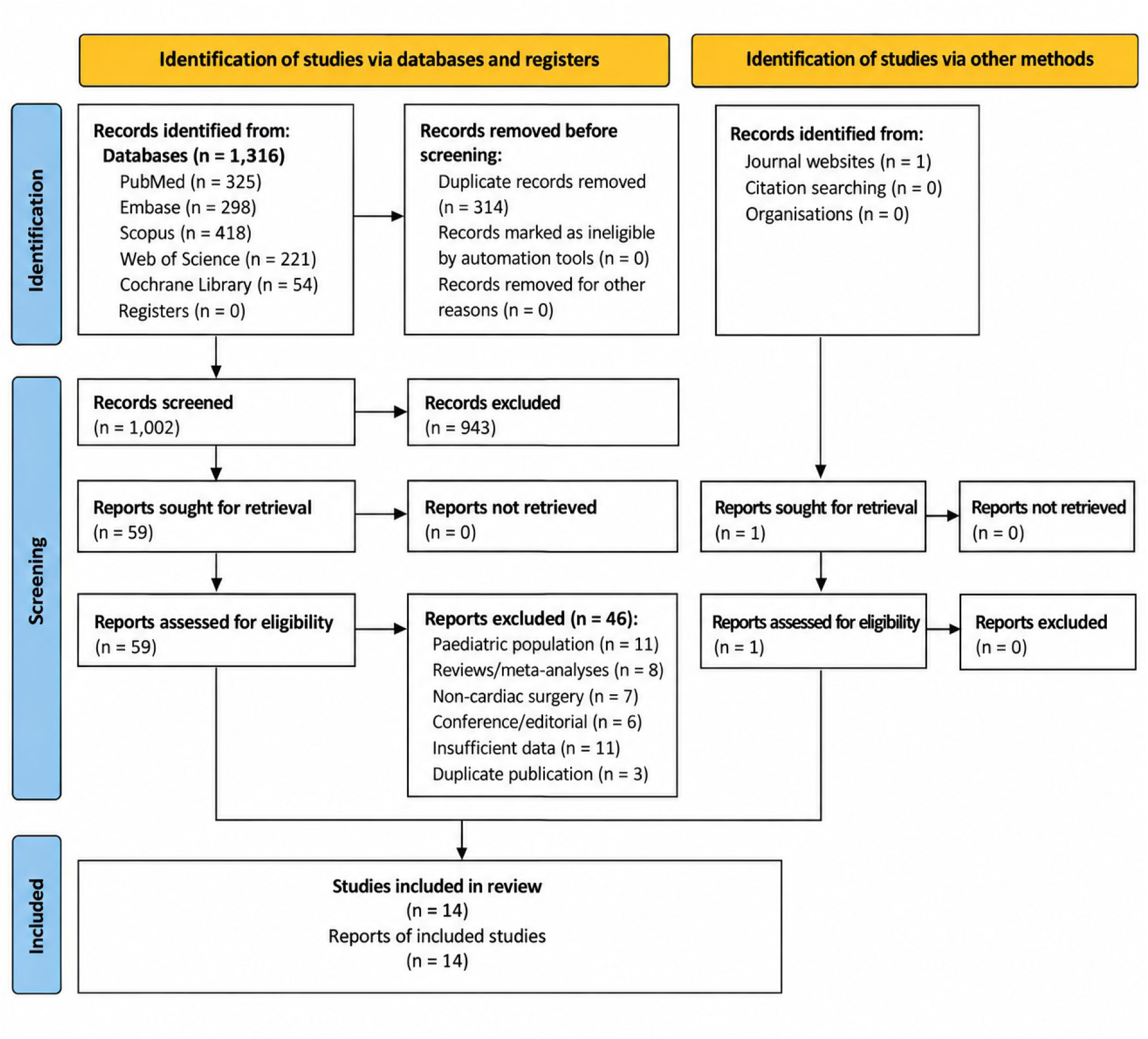
PRISMA 2020 flow diagram. Alt-text: PRISMA 2020 flow diagram showing 1,316 database records and one additional report identified through journal websites. After removal of 314 duplicates, 1,002 records were screened, 60 reports were assessed at full text, 46 were excluded, and 14 studies were included in the review and meta-analysis.

### 3.2 Characteristics of Included Studies

Table 1 summarizes the key characteristics of the studies included in the present analysis. Overall, 14 observational cohort studies comprising 74,571 patients were included. Early postoperative mortality and acute kidney injury or renal failure were evaluated in 8 studies each, while postoperative stroke and postoperative infection were reported in 4 studies each. Red blood cell transfusion was assessed in 9 studies. Propensity-score matching was used in 3 studies.

Among the 74,571 patients included in the 14 studies, 17,349 (23.3%) were classified as anaemic according to the criteria used in each study. The reported prevalence of anaemia ranged from 12.8% to 58.4%. Six studies used World Health Organization (WHO)-based criteria: Padmanabhan et al. (2019), Saracoglu et al. (2020), Tran et al. (2021), Hazen et al. (2022), Rubino et al. (2023), and Warner et al. (2024). Five studies used study-specific haemoglobin thresholds: Sanders et al. (2017), Tauriainen et al. (2017), Dai et al. (2018), Karkouti et al. (2018), and Al-Riyami et al. (2021). Haematocrit values were incorporated into the definition in two studies (Bangash et al., 2024; Shales et al., 2024), while Sain et al. (2024) additionally included ferritin levels. Overall, the studies used different criteria to define preoperative anaemia.

Among the nine studies reporting red blood cell transfusion, 8,805 of 14,405 anaemic patients (61.1%) received transfusions, compared with 14,562 of 51,087 non-anaemic patients (28.5%).

### 3.3 Primary Outcome

The primary outcome was in-hospital or 30-day mortality. Eight studies contributed to this analysis, including 59,769 patients and 772 reported deaths. Of these, 332 deaths occurred among anaemic patients and 440 among non-anaemic patients. Anaemic patients had significantly higher odds of mortality than non-anaemic patients (odds ratio [OR], 2.11; 95% confidence interval [CI], 1.40–3.17; I² = 58.4%; p = 0.0036) **(Figure 2).**

**Figure 2.**
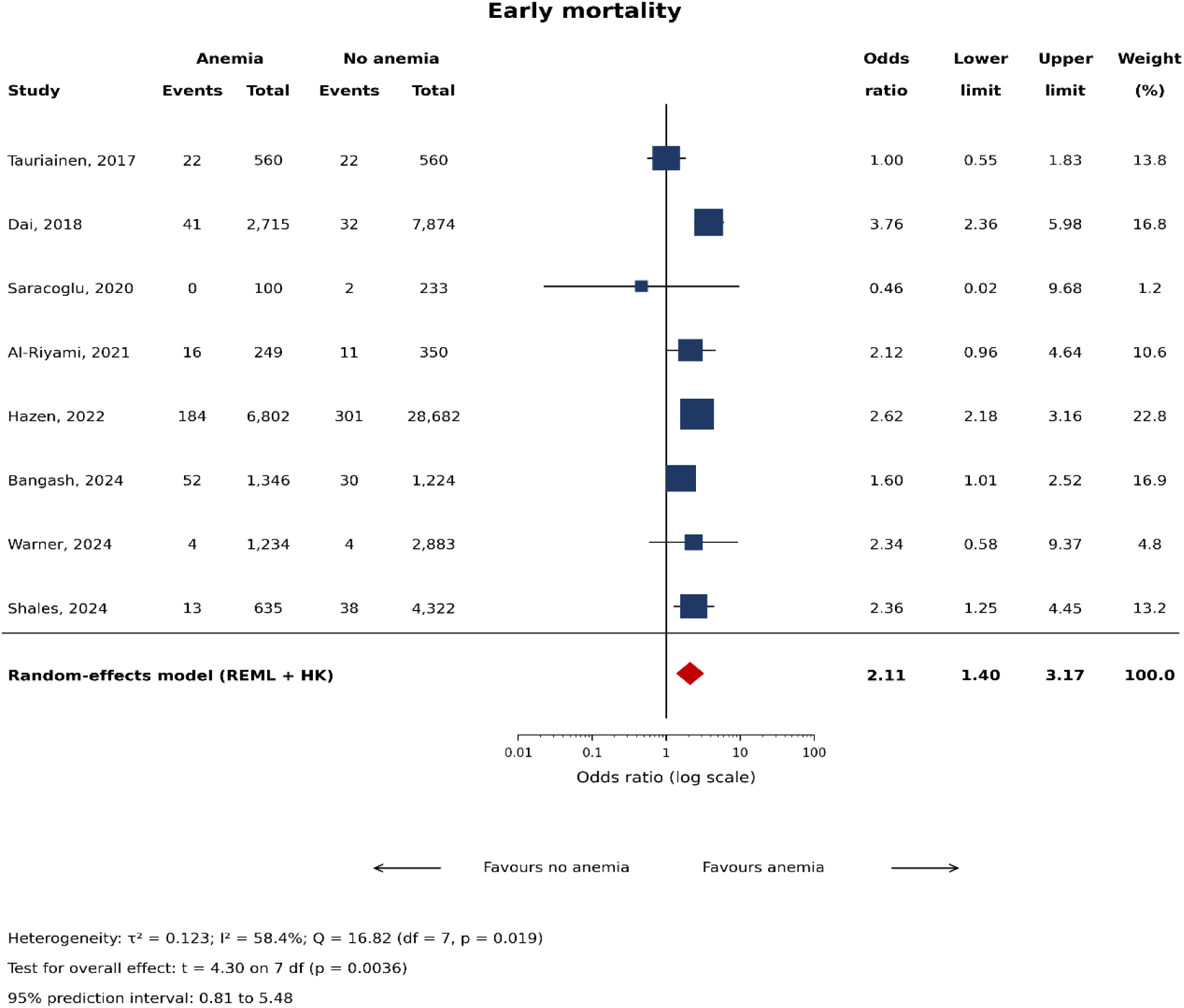
Forest plot for in-hospital or 30-day mortality. Odds ratios are shown with 95% confidence interval (CI). Alt-text: Forest plot of eight studies comparing in-hospital or 30-day mortality in anaemic versus non-anaemic adults undergoing cardiac surgery. The pooled random-effects odds ratio is 2.11 (95% CI 1.40–3.17; I² = 58.4%), indicating higher early mortality among anaemic patients

### 3.4 Secondary Outcomes

#### Acute Kidney Injury or Renal Failure

Eight studies, including 57,681 patients, evaluated acute kidney injury or renal failure. Preoperative anaemia was associated with higher odds of acute kidney injury or renal failure (OR, 2.43; 95% CI, 1.84–3.21; I² = 70.1%; p < 0.001) (Figure 3).

**Figure 3.**
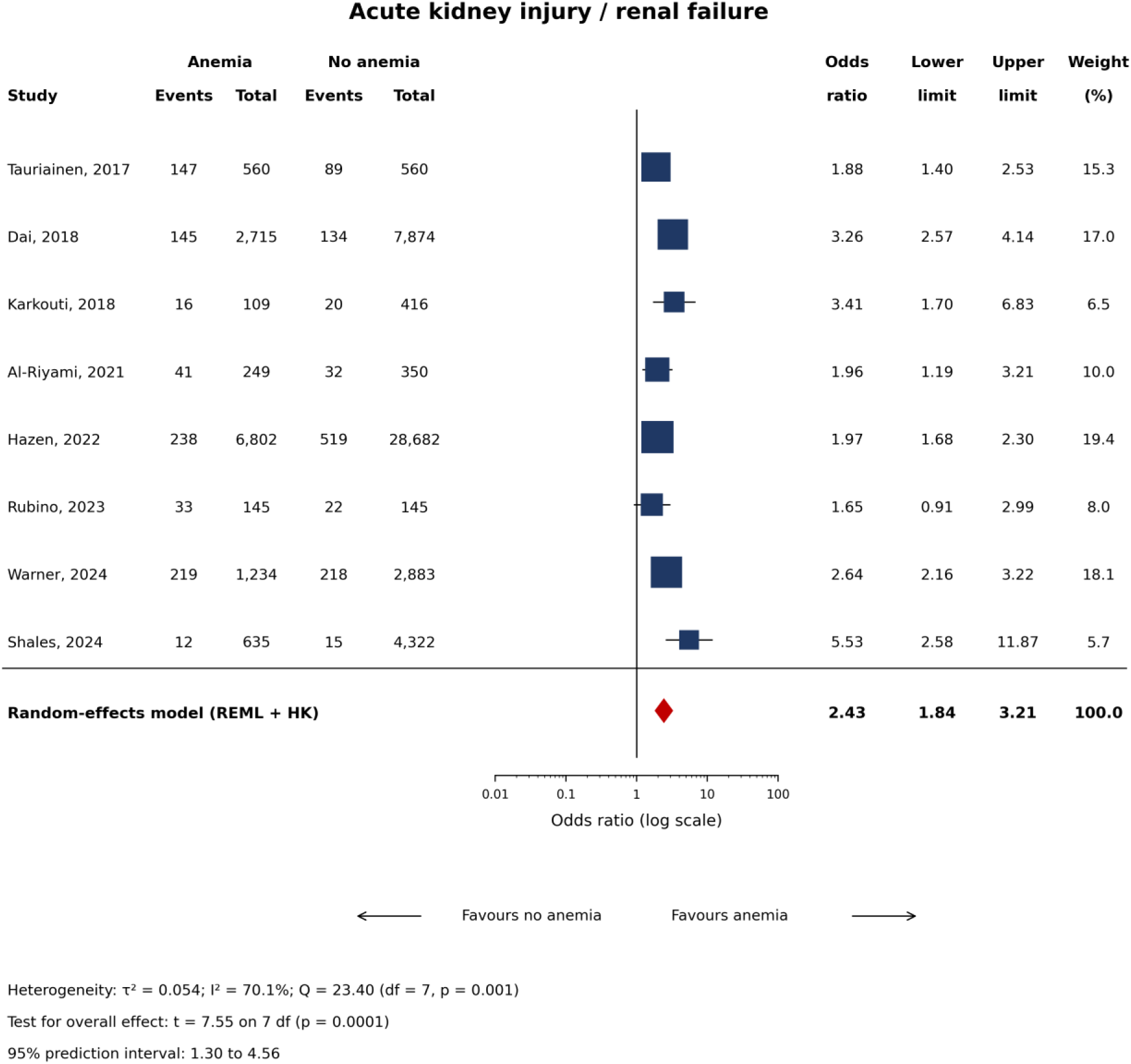
Forest plot for acute kidney injury or renal failure. Alt-text: Forest plot of eight studies comparing acute kidney injury or renal failure in anaemic versus non-anaemic adults undergoing cardiac surgery. The pooled random-effects odds ratio is 2.43 (95% CI 1.84–3.21; I² = 70.1%), indicating higher odds among anaemic patients.

#### Postoperative Stroke

Four studies involving 39,464 patients reported postoperative stroke. The odds of stroke were not significantly different between anaemic and non-anaemic patients (OR, 1.16; 95% CI, 0.43–3.17; I² = 56.3%; p = 0.6617) (Figure 4).

**Figure 4.**
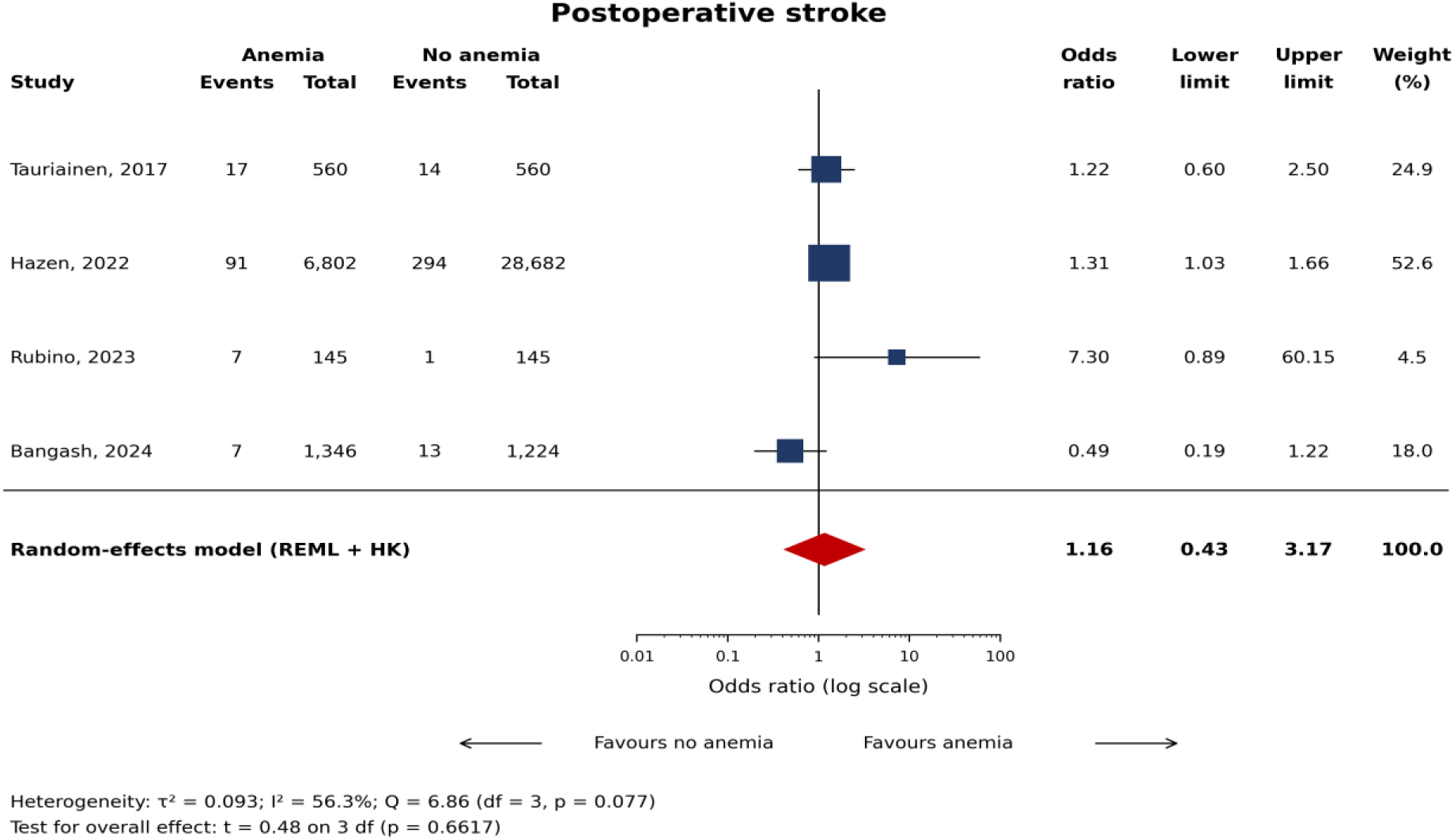
Forest plot for postoperative stroke. Alt-text: Forest plot of four studies comparing postoperative stroke in anaemic versus non-anaemic adults undergoing cardiac surgery. The pooled random-effects odds ratio is 1.16 (95% CI 0.43–3.17; I² = 56.3%), with no statistically significant difference between groups.

#### Postoperative Infection

Four studies involving 42,160 patients reported postoperative infection. Preoperative anaemia was associated with higher odds of postoperative infection (OR, 1.49; 95% CI, 1.21–1.82; I² = 0.0%; p = 0.0086) (Figure 5).

**Figure 5.**
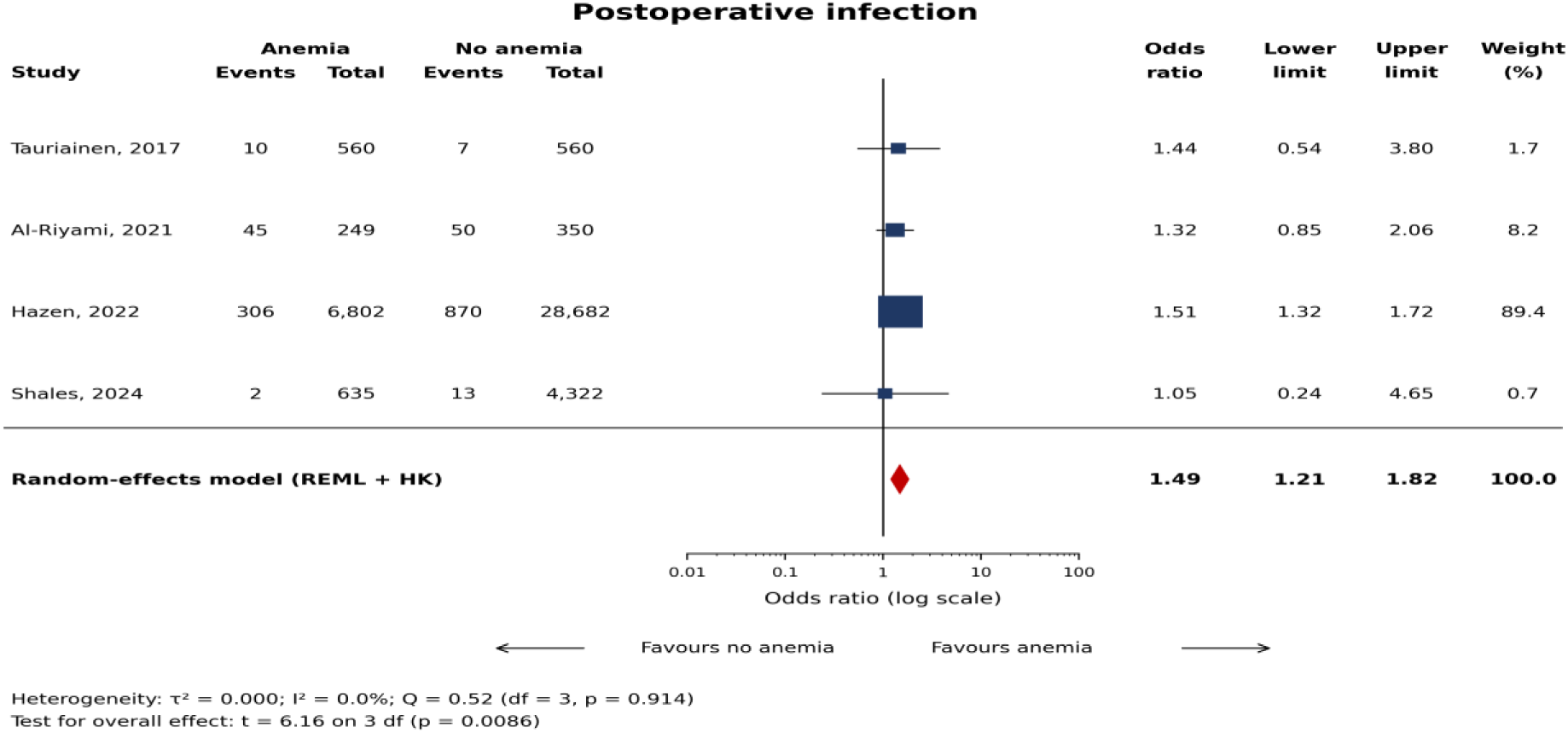
Forest plot for postoperative infection. Alt-text: Forest plot of four studies comparing postoperative infection in anaemic versus non-anaemic adults undergoing cardiac surgery. The pooled random-effects odds ratio is 1.49 (95% CI 1.21–1.82; I² = 0.0%), indicating higher odds of infection among anaemic patients.

#### Red Blood Cell Transfusion

Nine studies comprising 65,492 patients reported red blood cell transfusion. Among anaemic patients, 8,805 of 14,405 (61.1%) received a transfusion, compared with 14,562 of 51,087 (28.5%) among non-anaemic patients. Preoperative anaemia was associated with higher odds of red blood cell transfusion (OR, 3.66; 95% CI, 2.14–6.24; I² = 97.7%; p < 0.001) (Figure 6).

**Figure 6.**
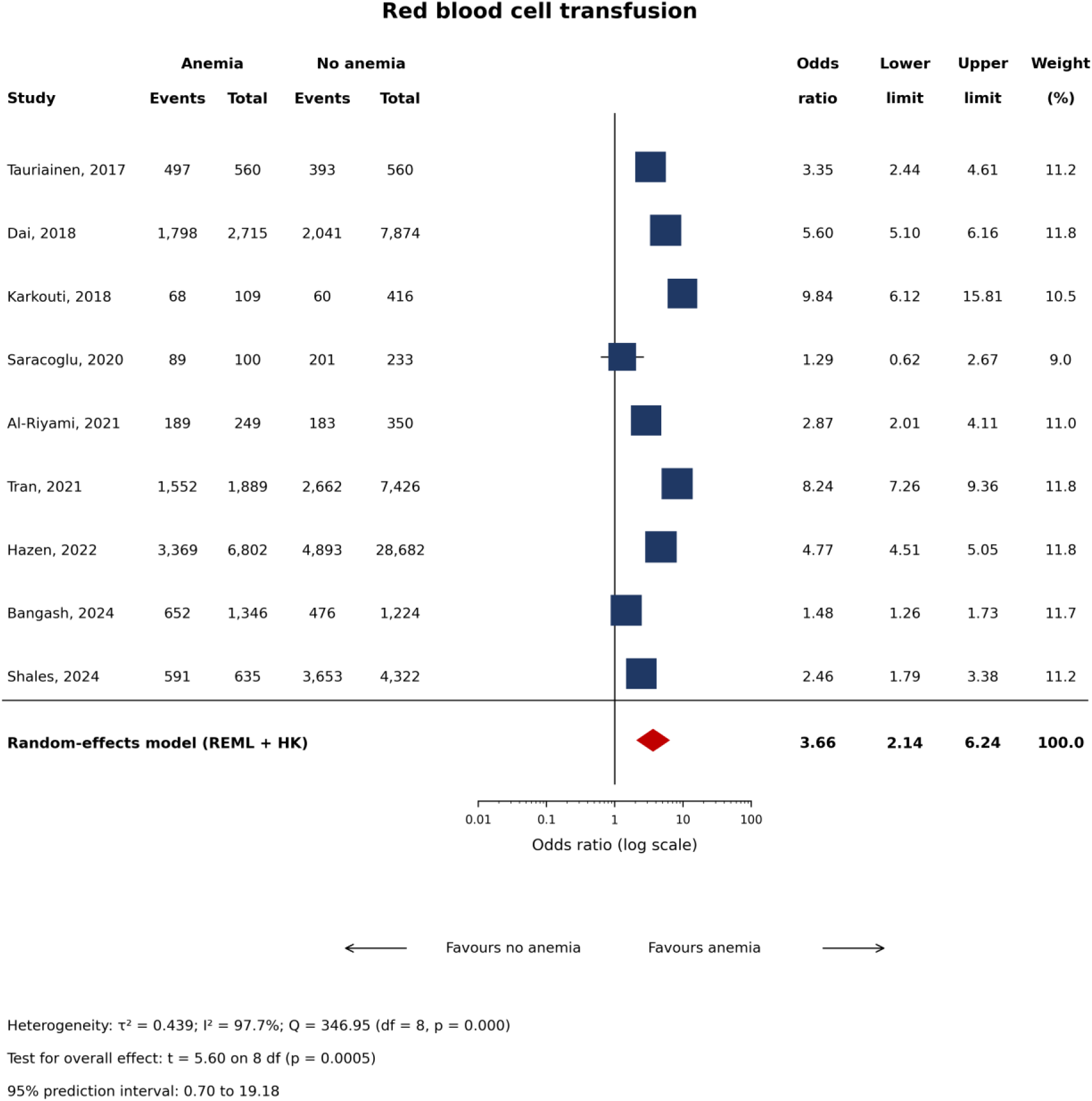
Forest plot for perioperative red-cell transfusion. Alt-text: Forest plot of nine studies comparing perioperative red-cell transfusion in anaemic versus non-anaemic adults undergoing cardiac surgery. The pooled random-effects odds ratio is 3.66 (95% CI 2.14–6.24; I² = 97.7%), indicating higher transfusion odds among anaemic patients with substantial between-study heterogeneity.

### 3.5 Subgroup and Sensitivity Analyses

Sensitivity analyses generally supported the primary findings. Associations with red blood cell transfusion, acute kidney injury or renal failure, and early mortality remained statistically significant across leave-one-out analyses. The association with postoperative infection lost statistical significance after excluding Hazen et al., whereas postoperative stroke remained non-significant. Excluding propensity-matched cohorts did not materially change the results for four outcomes; the mortality estimate increased to OR 2.43 (95% CI, 1.74–3.39), with heterogeneity decreasing from I² = 58.4% to 27.5%. For postoperative stroke, only two studies remained after exclusion, rendering the estimate uninterpretable. Restriction to studies with Newcastle–Ottawa Scale scores of 8–9 yielded broadly similar estimates. Detailed results are presented in **Supplementary eTable S8.**

Subgroup analyses showed a significant difference in early mortality according to surgery type (p for interaction = 0.0015). For red blood cell transfusion, significant subgroup differences were observed according to surgery type, anaemia definition, and transfusion timing (all p < 0.001). For acute kidney injury or renal failure, significant differences were observed according to anaemia definition, outcome definition and/or timing, study design, and geographic region. These findings should be interpreted with caution, as several subgroups included only a small number of studies. Subgroup analyses for red blood cell transfusion are presented in Figure 7.

**Figure 7.**
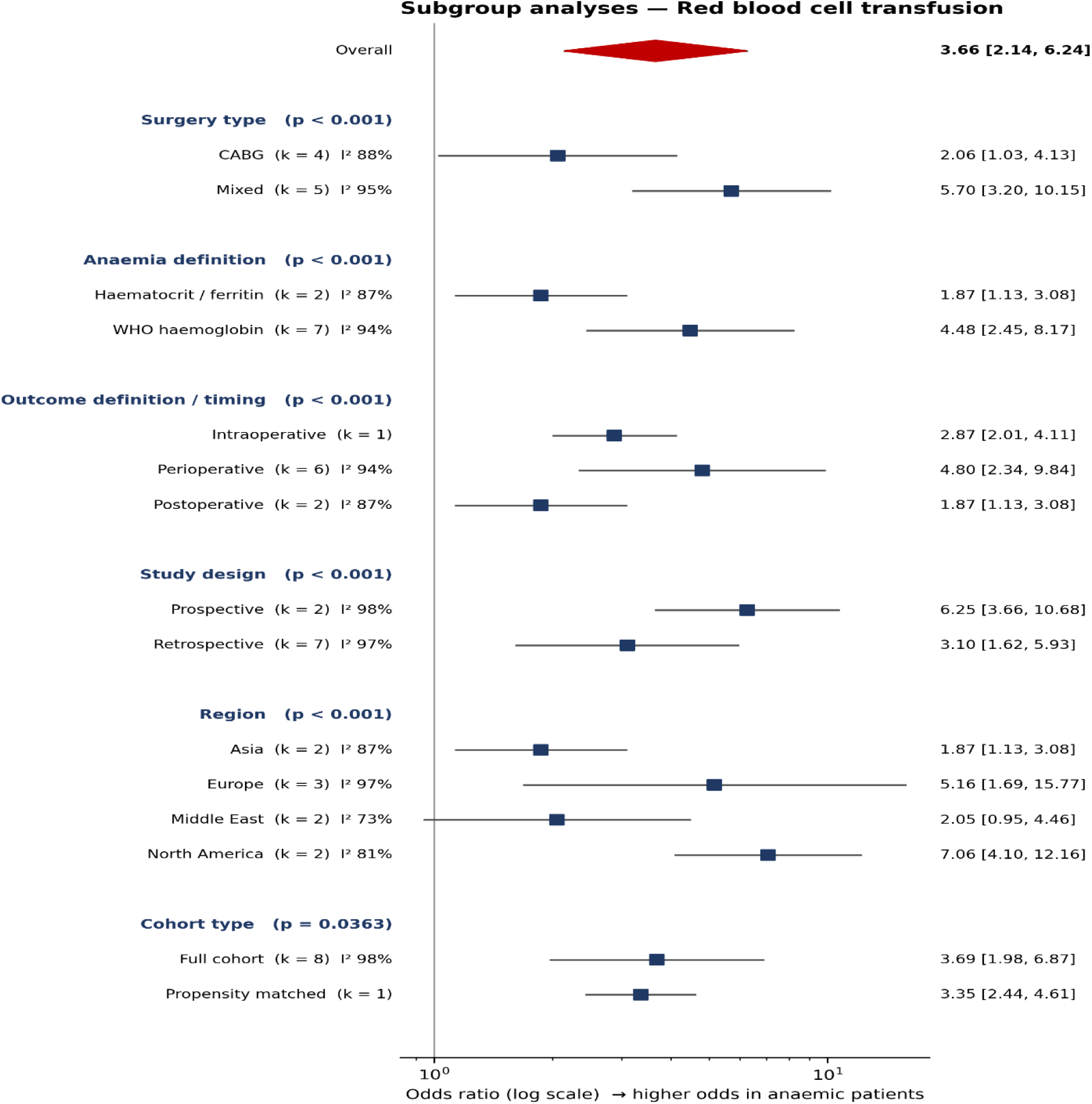
Subgroup analyses for red blood cell transfusion. Alt-text: Subgroup Forest plot for red blood cell transfusion across nine studies. The overall odds ratio is 3.66 (95% CI 2.14–6.24), with subgroup estimates displayed by surgery type, anaemia definition, outcome timing, study design, geographic region, and cohort type; several subgroups contain few studies.

### 3.6 Certainty of the Evidence

All 14 studies scored 7–9 on the Newcastle–Ottawa Scale. Agreement was observed for 122 of 126 domain ratings (96.8%), with a Cohen’s kappa of 0.784. However, the Newcastle–Ottawa summary score does not establish that confounding has been adequately controlled; therefore, the risk of bias was rated as serious for all outcomes under the GRADE framework.

Certainty was rated as low for acute kidney injury or renal failure and very low for red-cell transfusion, early mortality, postoperative infection, and postoperative stroke. Downgrading reflected the observational design, substantial or extreme heterogeneity, imprecision, non-equivalent outcome definitions, and sensitivity to influential studies, as detailed in Table 2.

**Table 2.** Summary of findings and certainty of evidence.

| Outcome | Assumed risk (non-anaemic) | Corresponding risk (anaemic) | Absolute difference | Relative effect OR (95% CI) | GRADE |
| --- | --- | --- | --- | --- | --- |
| Acute kidney injury / renal failure | 23/1,000 | 55/1,000 (42–71) | 31 more/1,000 (19–48 more) | 2.43 (1.84–3.21) | Low |
| Red blood cell transfusion | 285/1,000 | 593/1,000 (461–713) | 308 more/1,000 (176–428 more) | 3.66 (2.14–6.24) | Very low |
| Early mortality | 10/1,000 | 20/1,000 (13–30) | 10 more/1,000 (4–20 more) | 2.11 (1.40–3.17) | Very low |
| Postoperative infection | 28/1,000 | 41/1,000 (33–49) | 13 more/1,000 (5–21 more) | 1.49 (1.21–1.82) | Very low |
| Postoperative stroke | 11/1,000 | 12/1,000 (5–33) | 2 more/1,000 (6 fewer–22 more) | 1.16 (0.43–3.17) | Very low |

**Note:** Absolute risk estimates are descriptive translations of the pooled odds ratios based on the observed non-anaemic comparator risks and should not be interpreted as causal predictions. Baseline risks varied across cohorts, and residual confounding cannot be excluded.

### 3.7 Publication Bias

Publication bias was not formally assessed because none of the outcomes included 10 or more study estimates, which is generally considered the minimum number for meaningful assessment using funnel plots and Egger’s test. Therefore, publication bias was considered not assessable rather than assumed to be absent.

### 3.8 Adjusted Analyses

Adjusted estimates remained substantial for acute kidney injury (OR, 2.09; 95% CI, 1.66–2.64) and red blood cell transfusion (OR, 3.21; 95% CI, 1.95–5.28), whereas the association with early mortality was attenuated and no longer statistically significant (OR, 1.42; 95% CI, 0.59–3.40). Detailed adjusted analyses are presented in Supplementary eTable S9.

## 4. DISCUSSION

This updated synthesis indicates that preoperative anaemia is best understood as a clinically informative perioperative risk marker rather than a single, uniformly causal exposure. The primary analysis showed the strongest association for red-cell transfusion (OR 3.66), a substantial association for acute kidney injury or renal failure (OR 2.43), and a smaller association for early mortality (OR 2.11). These estimates should not be read as interchangeable measures of one biological effect. Transfusion is partly determined by clinician behaviour and institutional thresholds; renal injury reflects both perioperative oxygen delivery and baseline susceptibility; and mortality is a distal outcome with many competing causes.

### 4.1 Red-cell transfusion

The transfusion association was large but highly heterogeneous. The pooled odds ratio was 3.66, while study-specific estimates ranged from 1.29 to 9.84 and the prediction interval was 0.70–19.18, crossing the null and indicating substantial uncertainty about the expected effect in a new setting. All nine studies showed an association in the same direction; however, the magnitude was unlikely to be transportable across institutions. The non-anaemic comparator transfusion rate varied widely, from 14.4% to 86.3%, and the reported odds ratio was generally lower in studies with higher baseline transfusion rates. This pattern should be interpreted descriptively, because odds ratios are influenced by baseline risk and do not necessarily indicate true effect modification. The adjusted synthesis remained substantial (OR, 3.21; 95% CI, 1.95–5.28), supporting persistence of the association after adjustment for measured covariates, although residual confounding cannot be excluded. These findings are consistent with contemporary cardiac-surgery evidence linking preoperative anaemia to greater transfusion exposure (Groenewegen et al., 2026). Evidence from systematic reviews also suggests that perioperative intravenous iron may reduce transfusion requirements, although the comparative effectiveness of available anaemia-management strategies remains uncertain (Lau et al., 2024; Wang et al., 2025). Overall, the findings support systematic preoperative anaemia assessment and patient-blood-management strategies, but the pooled estimate should not be interpreted as a universal prediction rule or as evidence that anaemia alone determines the need for transfusion.

### 4.2 Acute kidney injury

The acute-kidney-injury association was directionally consistent across studies. Eight studies contributed to the primary model, all point estimates indicated higher risk among anaemic patients, and the prediction interval was 1.30–4.56. The pooled estimate was 2.43 (95% CI, 1.84–3.21; I² = 70.1%) and remained elevated and statistically significant in the adjusted synthesis (OR, 2.09; 95% CI, 1.66–2.64). Excluding matched cohorts increased the estimate to 2.67 (95% CI, 1.89–3.78). These findings are consistent with previous cardiac-surgery evidence linking preoperative anaemia with postoperative acute kidney injury (Karkouti et al., 2018; Warner et al., 2024; Padmanabhan et al., 2019), although the observational nature of the evidence does not establish that correcting anaemia will prevent renal injury.

### 4.3 Early mortality

The eight-study model yielded an odds ratio of 2.11 (95% CI, 1.40–3.17), but the prediction interval crossed the null (0.81–5.48), and the adjusted synthesis was attenuated to 1.42 (95% CI, 0.59–3.40) and was no longer statistically significant. Excluding the matched cohort strengthened the estimate to 2.43 (95% CI, 1.74–3.39) and reduced I² from 58.4% to 27.5%, suggesting that study design or cohort composition contributed to the observed heterogeneity. Taken together, this pattern is compatible with preoperative anaemia partly reflecting underlying comorbidity and overall perioperative vulnerability, rather than representing an independent causal determinant of mortality. This interpretation is consistent with previous cardiac-surgery evidence linking preoperative anaemia to mortality and other adverse outcomes (Padmanabhan et al., 2019; Ripoll et al., 2021; Warner et al., 2024), while the observational nature of the evidence limits causal inference. Therefore, the findings support enhanced perioperative risk assessment but do not establish that correcting anaemia alone will reduce early mortality.

### 4.4 Postoperative infection

Postoperative infection was pooled in the final analysis across four studies, yielding an odds ratio of 1.49 (95% CI, 1.21–1.82; I² = 0.0%). Two studies reported a composite infection endpoint, whereas two reported deep sternal wound infection alone. The numerical association is therefore not directly transportable to postoperative infection as a single harmonised endpoint, and certainty was downgraded for indirectness. This finding is consistent with previous cardiac-surgery evidence linking preoperative anaemia with postoperative infection, although the reported infection outcomes and their definitions have varied across studies (Padmanabhan et al., 2019). The estimate was sensitive to the largest contributing study; therefore, the narrow confidence interval should not be interpreted as evidence of high certainty, particularly given the limited number of studies, outcome heterogeneity, and observational study designs (Balshem et al., 2011).

### 4.5 Stroke

Stroke remained uninformative rather than null. Four studies contributed, with estimates ranging from 0.49 to 7.30 and a pooled random-effects odds ratio of 1.16 (95% CI, 0.43–3.17). The Mantel–Haenszel and Peto estimates were statistically positive (1.26 and 1.28, respectively), whereas the random-effects estimate was not; this divergence reflects between-study variance and should not be resolved by selecting the estimator that reaches significance. Excluding matched cohorts reversed the direction to 0.89 (95% CI, 0.34–2.29), but only two studies remained. Previous cardiac-surgery evidence has reported an association between preoperative anaemia and postoperative stroke, but the present analysis does not provide sufficiently precise or consistent evidence to confirm such an association (Padmanabhan et al., 2019).

### 4.6 Comparison with Previous Studies

Compared with the earlier cardiovascular surgery meta-analysis by Padmanabhan et al. (2019), the present review includes more recent cohort evidence and provides a clearer distinction between crude and adjusted estimates. This distinction is important when interpreting the findings, as crude estimates reflect the overall risk profile of patients with preoperative anaemia, whereas adjusted estimates account for measured differences in baseline characteristics and therefore provide a different perspective on the association between anaemia and postoperative outcomes. Because the adjustment variables, matching methods, and outcome definitions varied across studies, crude and adjusted estimates were considered separately rather than combined. The present review also highlights that a high Newcastle–Ottawa Scale score should not be interpreted as evidence that confounding has been adequately controlled, as adequate selection and outcome ascertainment do not necessarily eliminate residual confounding.

### 4.7 Clinical Implications

These findings support routine preoperative haemoglobin assessment and evaluation of potentially reversible causes of anaemia as part of perioperative optimisation. However, the available evidence does not establish that increasing haemoglobin to a specific threshold would reduce mortality, acute kidney injury or renal failure, postoperative infection, postoperative stroke, or the need for red blood cell transfusion. Anaemia management should therefore be individualised and integrated with patient blood-management strategies rather than guided by a single haemoglobin target. Prospective interventional studies are needed to determine whether defined preoperative anaemia-management pathways, including assessment and treatment of iron deficiency when appropriate, can improve patient-important outcomes.

### 4.8 Overall Interpretation

Overall, preoperative anaemia appears to be an important marker of heightened perioperative risk, but the evidence does not establish a uniform independent causal effect. The most consistent association was observed for acute kidney injury or renal failure, while red blood cell transfusion showed the largest but most heterogeneous association. The mortality association may be influenced by residual confounding, infection warrants cautious interpretation, and stroke remains inconclusive. Certainty of evidence was low for acute kidney injury or renal failure and very low for the other outcomes.

### 4.9 Limitations

Several limitations should be considered. All included studies were observational, so residual confounding and alternative explanations for the observed associations cannot be excluded. The primary synthesis pooled crude event counts, whereas published adjusted estimates were based on non-uniform covariate sets and could not be directly harmonised. Propensity-matched sub-cohorts were intentionally retained in the primary exact-count analysis, resulting in a mixture of matched and unmatched two-by-two evidence; matching-induced dependence may affect variance and weighting, making the exclusion of matched cohorts an important sensitivity analysis. Definitions of anaemia, acute kidney injury, infection, and mortality varied across studies, contributing to clinical and methodological heterogeneity. Small-study effects could not be reliably assessed because no outcome included ten or more estimates. In addition, the search was restricted to five databases and English-language publications, so relevant evidence may have been missed. Finally, length of stay and duration of mechanical ventilation could not be pooled because the reported summary statistics were incompatible or unavailable.

## 5. CONCLUSIONS

Preoperative anaemia is associated with clinically important postoperative outcomes in adult cardiac surgery, especially red-cell transfusion and acute kidney injury. The mortality and infection associations are less secure after accounting for confounding and endpoint variation, and the evidence for stroke is uninformative. Preoperative haemoglobin assessment and patient blood-management pathways are reasonable clinical priorities, while causal benefit from anaemia correction should be tested in prospective comparative studies.

## 6. Data availability and reproducibility

The analysis dataset, forest plots, PRISMA flow diagram, GRADE evidence profile, Summary of Findings table, quality assessment workbook, protocol decision log, and study contribution log are available as supplementary files accompanying this manuscript.

## Acknowledgements

None

