## Supplementary Material for "Association Between Preoperative Anemia and Postoperative Outcomes in Adults Undergoing Cardiac Surgery: A Systematic Review and Meta-analysis"

**TABLE OF CONTENTS**

| eTable S 1 | Preferred Reporting Items for Systematic Reviews and Meta-Analyses checklist |
| --- | --- |
| eTable S2a-e | Search strategy for databases |
| eTable S3 | Data extraction template |
| eTable S4 | Characteristics of included studies |
| eTable S5 | Final adjudicated Newcastle–Ottawa Scale assessments |
| eTable S6 | GRADE certainty assessment |
| eTable S7 | Pooled results |
| eTable S8 | Sensitivity analyses |
| eTable S9 | Alternative meta-analytic estimates using Mantel–Haenszel and Peto methods. |
| eTable S10 | Data Inventory |
| eFigure S1 | PRISMA 2020 flow diagram |
| eFigure S2 | Subgroup Forest plot for acute kidney injury or renal failure |
| eFigure S3 | Subgroup Forest plot for early mortality |
| eFigure S4 | Subgroup Forest plot for postoperative infection |
| eFigure S5 | Subgroup Forest plot for postoperative stroke |
| eFigure S**6** | Baujat plots for heterogeneity contribution |
| eFigure S**7** | Externally standardised residuals for the five postoperative outcomes |
| eFigure S**8** | Baseline transfusion rate |

**eTable S1:** Preferred Reporting Items for Systematic Reviews and Meta-Analyses checklist

| **Section and Topic** | **Item #** | **Checklist item** | **Location where item is reported** |
| --- | --- | --- | --- |
| **TITLE** | | |  |
| Title | 1 | Identify the report as a systematic review. | Page 1 |
| **ABSTRACT** | | |  |
| Abstract | 2 | See the PRISMA 2020 for Abstracts checklist. | Page 1 |
| **INTRODUCTION** | | |  |
| Rationale | 3 | Describe the rationale for the review in the context of existing knowledge. | Page 1 |
| Objectives | 4 | Provide an explicit statement of the objective(s) or question(s) the review addresses. | Page 2 |
| **METHODS** | | |  |
| Eligibility criteria | 5 | Specify the inclusion and exclusion criteria for the review and how studies were grouped for the syntheses. | Pages 3–5 |
| Information sources | 6 | Specify all databases, registers, websites, organisations, reference lists and other sources searched or consulted to identify studies. Specify the date when each source was last searched or consulted. | Page 3 |
| Search strategy | 7 | Present the full search strategies for all databases, registers and websites, including any filters and limits used. | Page 3 |
| Selection process | 8 | Specify the methods used to decide whether a study met the inclusion criteria, including how many reviewers screened each record and report, whether they worked independently, and any automation tools used. | Page 3 |
| Data collection process | 9 | Specify the methods used to collect data from reports, including how many reviewers collected data, whether they worked independently, and any processes for obtaining or confirming data. | Page 4 |
| Data items | 10a | List and define all outcomes for which data were sought and specify the methods used to decide which results to collect. | Pages 3–5; eTable S3, pp. 9–10 |
|  | 10b | List and define all other variables for which data were sought and describe any assumptions made about missing or unclear information. | Page 4; eTable S3, pp. 9–10 |
| Study risk of bias assessment | 11 | Specify the methods used to assess risk of bias in the included studies, including the tool(s), number of reviewers, independence, and any automation tools. | Page 4; eTable S5, pp. 11–12 |
| Effect measures | 12 | Specify for each outcome the effect measure(s) used in the synthesis or presentation of results. | Page 4-5 |
| Synthesis methods | 13a | Describe the processes used to decide which studies were eligible for each synthesis. | Pages 3–5 |
|  | 13b | Describe any methods required to prepare the data for presentation or synthesis, such as handling missing summary statistics or data conversions. | Pages 4–5 |
|  | 13c | Describe any methods used to tabulate or visually display results of individual studies and syntheses. | Page 5; Figures 2–6 |
|  | 13d | Describe any methods used to synthesize results and provide a rationale for the choice(s), including models, heterogeneity methods, and software. | Page 5 |
|  | 13e | Describe any methods used to explore possible causes of heterogeneity among study results. | Page 5 |
|  | 13f | Describe any sensitivity analyses conducted to assess robustness of the synthesized results. | Page 5; eTable S8, pp. 14–15 |
| Reporting bias assessment | 14 | Describe any methods used to assess risk of bias due to missing results in a synthesis. | Page 5 |
| Certainty assessment | 15 | Describe any methods used to assess certainty (or confidence) in the body of evidence for an outcome. | Page 4; eTable S6, pp. 12–13 |
| **RESULTS** | | |  |
| Study selection | 16a | Describe the results of the search and selection process, from the number of records identified to the number of studies included, ideally using a flow diagram. | Page 6; Figure 1 |
|  | 16b | Cite studies that might appear to meet the inclusion criteria, but which were excluded, and explain why they were excluded. | Page 5- 6; Figure 1 |
| Study characteristics | 17 | Cite each included study and present its characteristics. | Pages 6–7; Table 1 |
| Risk of bias in studies | 18 | Present assessments of risk of bias for each included study. | Page 16; eTable 5 |
| Results of individual studies | 19 | For all outcomes, present for each study summary statistics for each group and an effect estimate and its precision, ideally using structured tables or plots. | Pages 7–14; Figures 2–6 |
| Results of syntheses | 20a | For each synthesis, briefly summarise the characteristics and risk of bias among contributing studies. | Pages 7–14 |
|  | 20b | Present results of all statistical syntheses, including summary estimates, precision, and measures of statistical heterogeneity. | Pages 7, 11, 14–17; Figures 2–7; Supplementary eTables S7–S9, pp. 13–15 |
|  | 20c | Present results of all investigations of possible causes of heterogeneity among study results. | Pages 14–16; Figure 7, p. 16; eFigures S2–S8, pp. 18–24 |
|  | 20d | Present results of all sensitivity analyses conducted to assess robustness of the synthesized results. | Pages 14–15; eTable S8, pp. 14–15 |
| Reporting biases | 21 | Present assessments of risk of bias due to missing results for each synthesis assessed. | Page 17 |
| Certainty of evidence | 22 | Present assessments of certainty (or confidence) in the body of evidence for each outcome assessed. | **Pages 16–17; Table 2, p. 17; Supplementary eTable S6, pp. 12–13** |
| **DISCUSSION** | | |  |
| Discussion | 23a | Provide a general interpretation of the results in the context of other evidence. | **Pages 18–20** |
|  | 23b | Discuss any limitations of the evidence included in the review. | **Pages 18–21; particularly p. 21** |
|  | 23c | Discuss any limitations of the review processes used. | Page 21 |
|  | 23d | Discuss implications of the results for practice, policy, and future research. | Page 20–21 |
| **OTHER INFORMATION** | | |  |
| Registration and protocol | 24a | Provide registration information for the review, including register name and registration number, or state that the review was not registered. | Page 3 |
|  | 24b | Indicate where the review protocol can be accessed, or state that a protocol was not prepared. | Page 3 -PROSPERO record |
|  | 24c | Describe and explain any amendments to information provided at registration or in the protocol. | Not applicable |
| Support | 25 | Describe sources of financial or non-financial support for the review and the role of funders or sponsors. | Not reported in current manuscript |
| Competing interests | 26 | Declare any competing interests of review authors. | Not reported in current manuscript |
| Availability of data, code and other materials | 27 | Report which materials are publicly available and where they can be found. | Page 21; Supplementary eTable S10, pp. 15–16 |

From: Page MJ, McKenzie JE, Bossuyt PM, Boutron I, Hoffmann TC, Mulrow CD, et al. The PRISMA 2020 statement: an updated guideline for reporting systematic reviews. BMJ. 2021;372:n71. doi:10.1136/bmj.n71

**eTable S2a: Search strategy for databases**

| **Search** | **Query** |
| --- | --- |
| **Overview** |  |
| Database | MEDLINE & PubMed Central |
| Platform | PubMed |
| Date of search | 20/07/2026 |
| Number of results | 325 |
| **Syntax guide** |  |
| [Title/Abstract] | Words in title or abstract |
| **Anaemia-associated search terms** | |
| #1 | "Preoperative Anemia"[Title/Abstract] |
| #2 | "Preoperative Anaemia"[Title/Abstract] |
| #3 | "Anemia"[Title/Abstract] |
| #4 | "Anaemia"[Title/Abstract] |
| #5 | #1 OR #2 OR #3 OR #4 |
| **Cardiac Surgery-associated search terms** | |
| #6 | "Cardiac Surgery"[Title/Abstract] |
| #7 | "Cardiac Surgical Procedures"[Title/Abstract] |
| #8 | "Coronary Artery Bypass Grafting"[Title/Abstract] |
| #9 | "CABG"[Title/Abstract] |
| #10 | "Valve Surgery"[Title/Abstract] |
| #11 | #6 OR #7 OR #8 OR #9 OR #10 |
| **Outcome-associated terms** | |
| #12 | "Mortality" |
| #13 | "Acute Kidney Injury" |
| #14 | "AKI" |
| #15 | "Stroke" |
| #16 | "Blood Transfusion" |
| #17 | "Postoperative Infection" |
| #18 | "Hospital Length of Stay" |
| #19 | "ICU Length of Stay" |
| #20 | "Postoperative Complications" |
| #21 | #12 OR #13 OR #14 OR #15 OR #16 OR #17 OR #18 OR #19 OR #20 |
| **Final search** |  |
| #22 | #5 AND #11 AND #21 |

**eTable S2b: Search strategy for databases**

| **Search** | **Query** |
| --- | --- |
| **Overview** |  |
| Database | Scopus |
| Platform | Scopus |
| Number of results | 418 |
| **Syntax guide** |  |
| TITLE-ABS-KEY | Title, Abstract, and Keywords |
| **Anaemia-associated search terms** | |
| #1 | TITLE-ABS-KEY ("Preoperative Anemia") |
| #2 | TITLE-ABS-KEY ("Preoperative Anaemia") |
| #3 | TITLE-ABS-KEY ("Anemia") |
| #4 | TITLE-ABS-KEY ("Anaemia") |
| #5 | #1 OR #2 OR #3 OR #4 |
| **Cardiac Surgery-associated search terms** | |
| #6 | TITLE-ABS ("Cardiac Surgery") |
| #7 | TITLE-ABS ("Cardiac Surgical Procedures") |
| #8 | TITLE-ABS ("Coronary Artery Bypass Grafting") |
| #9 | TITLE-ABS ("CABG") |
| #10 | TITLE-ABS ("Valve Surgery") |
| #11 | #6 OR #7 OR #8 OR #9 OR #10 |
| **Outcome-associated terms** | |
| #12 | TITLE-ABS ("Mortality") |
| #13 | TITLE-ABS ("Acute Kidney Injury") |
| #14 | TITLE-ABS ("AKI") |
| #15 | TITLE-ABS ("Stroke") |
| #16 | TITLE-ABS ("Blood Transfusion") |
| #17 | TITLE-ABS ("Postoperative Infection") |
| #18 | TITLE-ABS ("Hospital Length of Stay") |
| #19 | TITLE-ABS ("ICU Length of Stay") |
| #20 | TITLE-ABS ("Postoperative Complications") |
| #21 | #12 OR #13 OR #14 OR #15 OR #16 OR #17 OR #18 OR #19 OR #20 |
| **Final search** |  |
| #22 | #5 AND #11 AND #21 |

**eTable S2c: Search strategy for databases**

| **Search** | **Query** |
| --- | --- |
| Overview |  |
| Database | Cochrane Library |
| Platform | Cochrane Library |
| Number of results | 54 |
| Syntax guide |  |
| :ti,ab,kw | Title, Abstract, and Keywords |
| Anaemia-associated search terms | |
| #1 | "preoperative anemia":ti,ab,kw |
| #2 | "preoperative anaemia":ti,ab,kw |
| #3 | anemia:ti,ab,kw |
| #4 | anaemia:ti,ab,kw |
| #5 | #1 OR #2 OR #3 OR #4 |
| Cardiac Surgery-associated search terms | |
| #6 | "cardiac surgery":ti |
| #7 | "cardiac surgical procedures":ti |
| #8 | CABG:ti |
| #9 | "coronary artery bypass grafting":ti |
| #10 | "valve surgery":ti |
| #11 | #6 OR #7 OR #8 OR #9 OR #10 |
| Outcome-associated terms | |
| #12 | mortality:ti,ab,kw |
| #13 | "acute kidney injury":ti,ab,kw |
| #14 | AKI:ti,ab,kw |
| #15 | stroke:ti,ab,kw |
| #16 | "blood transfusion":ti,ab,kw |
| #17 | "postoperative infection":ti,ab,kw |
| #18 | "hospital length of stay":ti,ab,kw |
| #19 | "ICU length of stay":ti,ab,kw |
| #20 | "postoperative complications":ti,ab,kw |
| #21 | #12 OR #13 OR #14 OR #15 OR #16 OR #17 OR #18 OR #19 OR #20 |
| Final search |  |
| #22 | #5 AND #11 AND #21 |

**eTable S2d: Search strategy for databases**

| **Search** | **Query** |
| --- | --- |
| **Overview** |  |
| Database | Web of Science |
| Platform | Web of Science |
| Number of results | 221 |
| **Syntax guide** |  |
| TS= | Topic search |
| **Anaemia-associated search terms** | |
| #1 | TS=("preoperative anemia") |
| #2 | TS=("preoperative anaemia") |
| #3 | TS=(anemia) |
| #4 | TS=(anaemia) |
| #5 | #1 OR #2 OR #3 OR #4 |
| **Cardiac Surgery-associated search terms** | |
| #6 | TS=("cardiac surgery") |
| #7 | TS=("cardiac surgical procedures") |
| #8 | TS=(CABG) |
| #9 | TS=("coronary artery bypass grafting") |
| #10 | TS=("valve surgery") |
| #11 | #6 OR #7 OR #8 OR #9 OR #10 |
| **Outcome-associated terms** | |
| #12 | TS=(mortality) |
| #13 | TS=("acute kidney injury") |
| #14 | TS=(AKI) |
| #15 | TS=(stroke) |
| #16 | TS=("blood transfusion") |
| #17 | TS=("postoperative infection") |
| #18 | TS=("hospital length of stay") |
| #19 | TS=("ICU length of stay") |
| #20 | TS=("postoperative complications") |
| #21 | #12 OR #13 OR #14 OR #15 OR #16 OR #17 OR #18 OR #19 OR #20 |
| **Final search** |  |
| #22 | #5 AND #11 AND #21 |

**eTable S2e: Search strategy for databases**

| **Search** | **Query** |
| --- | --- |
| Overview |  |
| Database | Embase |
| Platform | Embase |
| Number of results | 298 |
| Anaemia-associated search terms | |
| #1 | "preoperative anemia" |
| #2 | "preoperative anaemia" |
| #3 | anemia |
| #4 | anaemia |
| #5 | #1 OR #2 OR #3 OR #4 |
| Cardiac Surgery-associated search terms | |
| #6 | "cardiac surgery" |
| #7 | "cardiac surgical procedures" |
| #8 | CABG |
| #9 | "coronary artery bypass grafting" |
| #10 | "valve surgery" |
| #11 | #6 OR #7 OR #8 OR #9 OR #10 |
| Outcome-associated terms | |
| #12 | mortality |
| #13 | "Acute kidney injury" |
| #14 | AKI |
| #15 | stroke |
| #16 | "blood transfusion" |
| #17 | "postoperative infection" |
| #18 | "hospital length of stay" |
| #19 | "intensive care unit length of stay" |
| #20 | "postoperative complications" |
| #21 | #12 OR #13 OR #14 OR #15 OR #16 OR #17 OR #18 OR #19 OR #20 |
| Final search |  |
| #22 | #5 AND #11 AND #21 |

**eTable S3: Data extraction template**

Study characteristics: Study ID, title, authors, publication, country, inclusion/exclusion criteria, dates.

Patient demographics: Sample size, age, sex, hemoglobin level, hematocrit, anemia definition, type of surgery, and relevant postoperative outcomes.

Details about surgery: Type of surgery (CABG, valve surgery, combined CABG/valve surgery, redo cardiac surgery, mixed cardiac surgery, or other eligible cardiac procedures).

Primary outcomes: mortality.

Secondary outcomes: infection, acute kidney injury, stroke, blood transfusion, hospital length of stay, ICU length of Stay, postoperative complications

**eTable S4: Characteristics of included studies**

| **Study** | **Country / centre** | **Anaemia definition** | **Surgery** | **Anaemic n (%)** | **Total N** | **Mortality** | **AKI** | **Stroke** | **Infection** | **Matched** |
| --- | --- | --- | --- | --- | --- | --- | --- | --- | --- | --- |
| Sanders et al., 2017 | UK (single-centre) | Hb <13 men; <12 women | Mixed | 139 (31.5%) | 442 | No | No | No | Yes | No |
| Tauriainen et al., 2017 | Finland (single-centre) | Hb <13 men; <12 women | CABG | 662 (24.0%) | 2,761 | Yes | Yes | Yes | Yes | Yes |
| Dai et al., 2018 | USA (single-centre) | Hb <13 men; <12 women | Mixed | 2,715 (26.0%) | 10,589 | Yes | Yes | No | No | No |
| Karkouti et al., 2018 | Canada (single-centre) | Hb <13 men; <12 women | Mixed | 109 (21.0%) | 525 | No | Yes | No | No | No |
| Padmanabhan et al., 2019 | UK (single-centre) | Hb <13 men; <12 women | Mixed | 1,170 (50.0%) | 2,340 | Yes | No | No | No | Yes |
| Saracoglu et al., 2020 | Türkiye (single-centre) | Hb <13 men; <12 women | CABG | 100 (30.0%) | 333 | Yes | No | Yes | No | No |
| Al-Riyami et al., 2021 | Oman (single-centre) | Hb <13 men; <12 women | Mixed | 249 (41.6%) | 599 | Yes | Yes | No | Yes | No |
| Tran et al., 2021 | Norway (single-centre) | Hb <13 men; <12 women | Mixed | 1,889 (20.3%) | 9,315 | Yes | No | No | No | No |
| Hazen et al., 2022 | Netherlands (multicentre) | Hb ≤12.9 men; ≤11.9 women | Mixed | 6,802 (19.2%) | 35,484 | Yes | Yes | Yes | Yes | No |
| Rubino et al., 2023 | Italy (single-centre) | WHO sex-specific criteria | Redo | 239 (58.4%) | 409 | No | Yes | Yes | No | Yes |
| Bangash et al., 2024 | Pakistan (single-centre) | Hct ≤33%; Hb <13 men; <12 women | CABG | ≈1,346 (52.37%) | 2,570 | Yes | No | Yes | No | No |
| Shales et al., 2024 | India (single-centre) | Hct <33% | OPCAB | 635 (12.8%) | 4,957 | Yes | Yes | No | Yes | No |
| Warner et al., 2024 | USA (single-centre) | Hb <13 men; <12 women | Mixed | 1,234 (30.0%) | 4,117 | Yes | Yes | No | No | No |
| Sain et al., 2024 | India (single-centre) | Hb <13 men; <12 women plus ferritin <30 ng/mL | Mixed | 60 (46.2%) | 130 | Yes | No | No | Yes | No |

Abbreviations: AKI, acute kidney injury; CABG, coronary artery bypass grafting; Hb, haemoglobin; Hct, haematocrit; OPCAB, off-pump coronary artery bypass; WHO, World Health Organization. Matched indicates a propensity-matched cohort where explicitly reported.

**eTable S5 — Final adjudicated Newcastle–Ottawa Scale assessments**

| **Final Adjudicated NOS Assessment (Newcastle–Ottawa Scale).** | | | | | | | | | | | | | |
| --- | --- | --- | --- | --- | --- | --- | --- | --- | --- | --- | --- | --- | --- |
| **Study** | **S1 Representativeness** | **S2 Selection of Non-Exposed Cohort** | **S3 Ascertainment of Exposure** | **S4 Outcome Absent at Start** | **Selection (0–4)** | **C1 Key Confounders** | **C2 Additional Confounders** | **Comparability (0–2)** | **O1 Assessment of Outcome** | **O2 Follow-up Long Enough** | **O3 Adequacy of Follow-up** | **Outcome (0–3)** | **Total NOS (0–9)** |
| Sanders et al., 2017 | 0 | 1 | 1 | 1 | ★★★☆ | 1 | 1 | ★★ | 1 | 1 | 1 | ★★★ | 8 |
| Tauriainen et al., 2017 | 1 | 1 | 1 | 1 | ★★★★ | 1 | 1 | ★★ | 1 | 1 | 1 | ★★★ | 9 |
| Dai et al., 2018 | 1 | 1 | 1 | 1 | ★★★★ | 0 | 0 | ☆☆ | 1 | 1 | 1 | ★★★ | 7 |
| Karkouti et al., 2018 | 0 | 1 | 1 | 1 | ★★★☆ | 1 | 1 | ★★ | 1 | 1 | 1 | ★★★ | 8 |
| Padmanabhan et al., 2019 | 1 | 1 | 1 | 1 | ★★★★ | 1 | 1 | ★★ | 1 | 1 | 1 | ★★★ | 9 |
| Saracoglu et al., 2020 | 1 | 1 | 1 | 1 | ★★★★ | 0 | 0 | ☆☆ | 1 | 1 | 1 | ★★★ | 7 |
| Al-Riyami et al., 2021 | 1 | 1 | 1 | 1 | ★★★★ | 1 | 1 | ★★ | 1 | 1 | 1 | ★★★ | 9 |
| Tran et al., 2021 | 0 | 1 | 1 | 1 | ★★★☆ | 1 | 1 | ★★ | 1 | 1 | 1 | ★★★ | 8 |
| Hazen et al., 2022 | 1 | 1 | 1 | 1 | ★★★★ | 1 | 1 | ★★ | 1 | 1 | 1 | ★★★ | 9 |
| Rubino et al., 2023 | 1 | 1 | 1 | 1 | ★★★★ | 1 | 1 | ★★ | 1 | 1 | 1 | ★★★ | 9 |
| Bangash et al., 2024 | 1 | 1 | 1 | 1 | ★★★★ | 0 | 0 | ☆☆ | 1 | 1 | 1 | ★★★ | 7 |
| Shales et al., 2024 | 1 | 1 | 1 | 1 | ★★★★ | 1 | 1 | ★★ | 1 | 1 | 1 | ★★★ | 9 |
| Warner et al., 2024 | 1 | 1 | 1 | 1 | ★★★★ | 1 | 1 | ★★ | 1 | 1 | 1 | ★★★ | 9 |
| Sain et al., 2024 | 0 | 1 | 1 | 1 | ★★★☆ | 1 | 1 | ★★ | 1 | 1 | 0 | ★★☆ | 7 |

**eTable S6. GRADE certainty assessment**

| **Outcome** | **Risk of bias** | **Inconsistency** | **Indirectness** | **Imprecision** | **Publication bias** | **Upgrade** | **Certainty** |
| --- | --- | --- | --- | --- | --- | --- | --- |
| Acute kidney injury | Serious −1 | Not serious | Not serious | Not serious | Not assessable | +1 large effect | **LOW** |
| Red cell transfusion | Serious −1 | Serious −1 | Not serious | Not serious | Not assessable | +1 large effect | **VERY LOW** |
| Early mortality | Serious −1 | Serious −1 | Not serious | Not serious | Not assessable | None | **VERY LOW** |
| Postoperative infection | Serious −1 | Not serious | Serious −1 | Not serious | Not assessable | None | **VERY LOW** |
| Postoperative stroke | Serious −1 | Serious −1 | Not serious | Very serious −2 | Not assessable | None | **VERY LOW** |

**eTable S7: Pooled results**

| **Outcome** | **Studies** | **Participants** | **Events**  anaemic | **Events**  comparator | **Pooled OR (95% CI)** | **p** | **I²** | **95% prediction interval** |
| --- | --- | --- | --- | --- | --- | --- | --- | --- |
| Red cell transfusion | 9 | 65,492 | 8,805 | 14,562 | 3.66 (2.14–6.24) | <0.001 | 97.7% | 0.70–19.18 |
| Acute kidney injury | 8 | 57,681 | 851 | 1,049 | 2.43 (1.84–3.21) | <0.001 | 70.1% | 1.30–4.56 |
| Early mortality | 8 | 59,769 | 332 | 440 | 2.11 (1.40–3.17) | 0.004 | 58.4% | 0.81–5.48 |
| Postoperative infection | 4 | 42,160 | 363 | 940 | 1.49 (1.21–1.82) | 0.009 | 0.0% | Not reported (k<5) |
| Postoperative stroke | 4 | 39,464 | 122 | 322 | 1.16 (0.43–3.17) | 0.66 | 56.3% | Not reported (k<5) |

**eTable S8. Sensitivity analyses.**

| **Outcome** | **Primary** | **Excluding matched cohorts** | **Restricted to NOS 8–9** | **Leave-one-out range** |
| --- | --- | --- | --- | --- |
| Transfusion | 3.66 (2.14–6.24) | 3.69 (1.98–6.87) | 4.57 (2.53–8.25) | 3.27–4.15, all significant |
| Acute kidney injury | 2.43 (1.84–3.21) | 2.67 (1.89–3.78) | 2.26 (1.70–2.99) | 2.26–2.56, all significant |
| Early mortality | 2.11 (1.40–3.17) | 2.43 (1.74–3.39), I² 27.5% | 2.01 (1.14–3.56) | 1.90–2.43, all significant |
| Infection | 1.49 (1.21–1.82) | 1.49 (1.12–1.96) | 1.49 (1.21–1.82) | 1.32–1.50, loses significance without Hazen |
| Stroke | 1.16 (0.43–3.17) | 0.89 (0.34–2.29) | 1.33 (0.76–2.31) | 1.06–1.33, never significant |

**eTable S9. Alternative meta-analytic estimates using Mantel–Haenszel and Peto methods.**

| **Outcome** | **Random effects (REML + HK)** | **Peto** | **Mantel-Haenszel** | **Adjusted estimates** |
| --- | --- | --- | --- | --- |
| Red cell transfusion | 3.66 (2.14–6.24) | 5.17 (4.95–5.39) | 4.69 (4.49–4.89) | 3.21 (1.95–5.28), k=5 |
| Acute kidney injury | 2.43 (1.84–3.21) | 2.61 (2.34–2.90) | 2.32 (2.10–2.55) | 2.09 (1.66–2.64), k=7 |
| Early mortality | 2.11 (1.40–3.17) | 2.77 (2.33–3.28) | 2.34 (2.01–2.71) | 1.42 (0.59–3.40), k=4 |
| Postoperative infection | 1.49 (1.21–1.82) | 1.55 (1.35–1.77) | 1.48 (1.31–1.68) | Not poolable |
| Postoperative stroke | 1.16 (0.43–3.17) | 1.28 (1.02–1.61) | 1.26 (1.02–1.57) | Not poolable |

eTable S10. Data Inventory

| **File** | **Purpose** |
| --- | --- |
| 01_meta_engine.py | Python meta-analysis engine and effect-size calculations |
| 02_analysis_R.R | R analysis, random-effects models, heterogeneity, and pooled results |
| 03_analysis_Stata.do | Independent Stata replication syntax |
| 04_influence_diagnostics.py | Leave-one-out and influence diagnostics |
| 05_figures.py | Forest-plot and figure generation |
| 06_grade_and_tables.py | GRADE evidence profile, Summary of Findings, and tables |
| 07_prisma_diagram.py | PRISMA 2020 diagram generation |
| README.md | Reproducibility instructions and file descriptions |
| anemia_binary.csv / anemia_adjusted.csv | Primary crude and secondary adjusted analysis datasets |
| anemia_binary.dta / anemia_adjusted.dta | Stata-format analysis datasets |

**eFigure S1. PRISMA 2020 flow diagram showing the identification, screening, eligibility assessment, and inclusion of studies.**

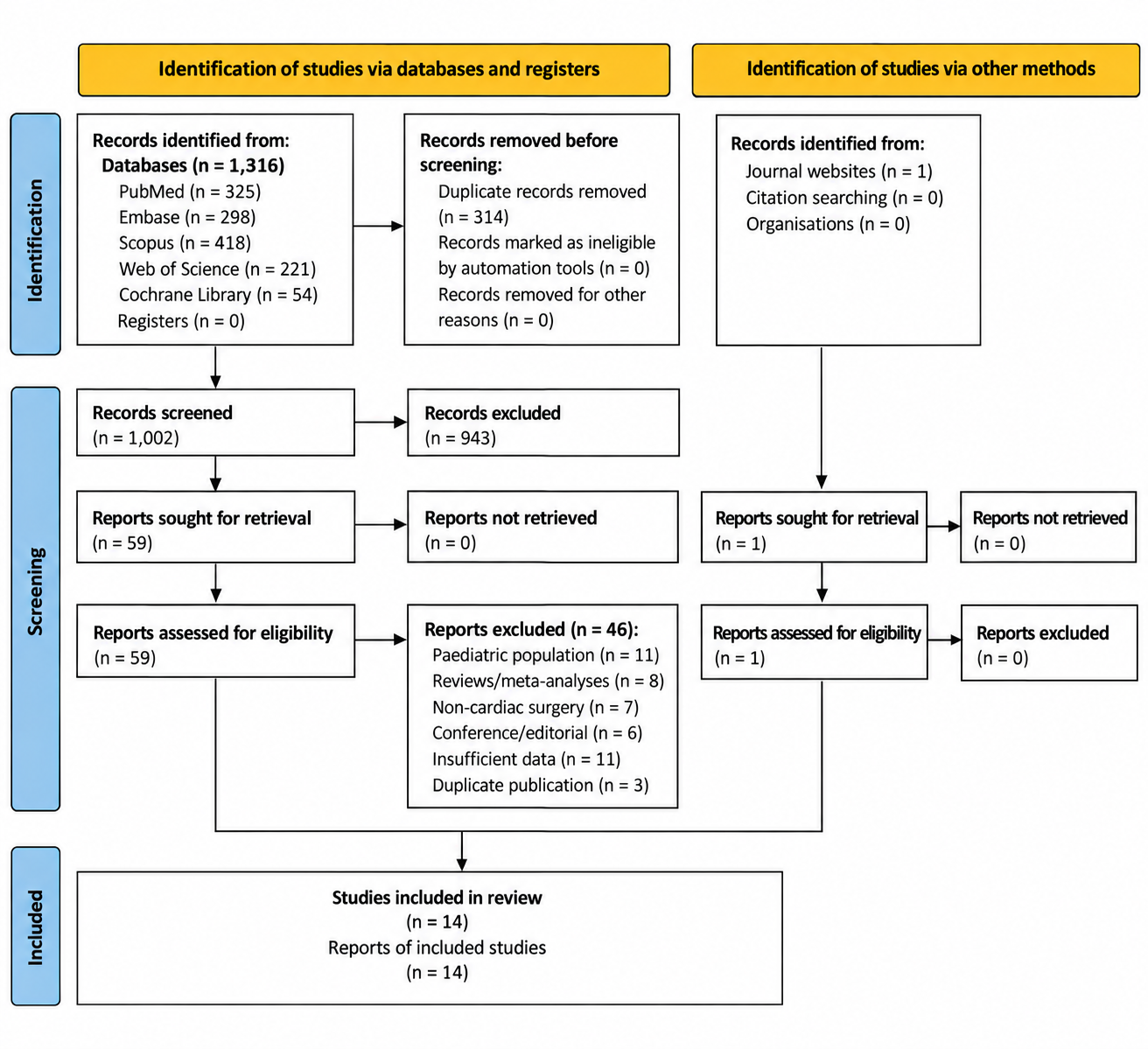

Alt-text: PRISMA 2020 flow diagram showing 1,316 database records and one additional report identified through journal websites. After removal of 314 duplicates, 1,002 records were screened, 60 reports were assessed at full text, 46 were excluded, and 14 studies were included in the review and meta-analysis.

**eFigure S2: Subgroup analysis for acute kidney injury or renal failure**

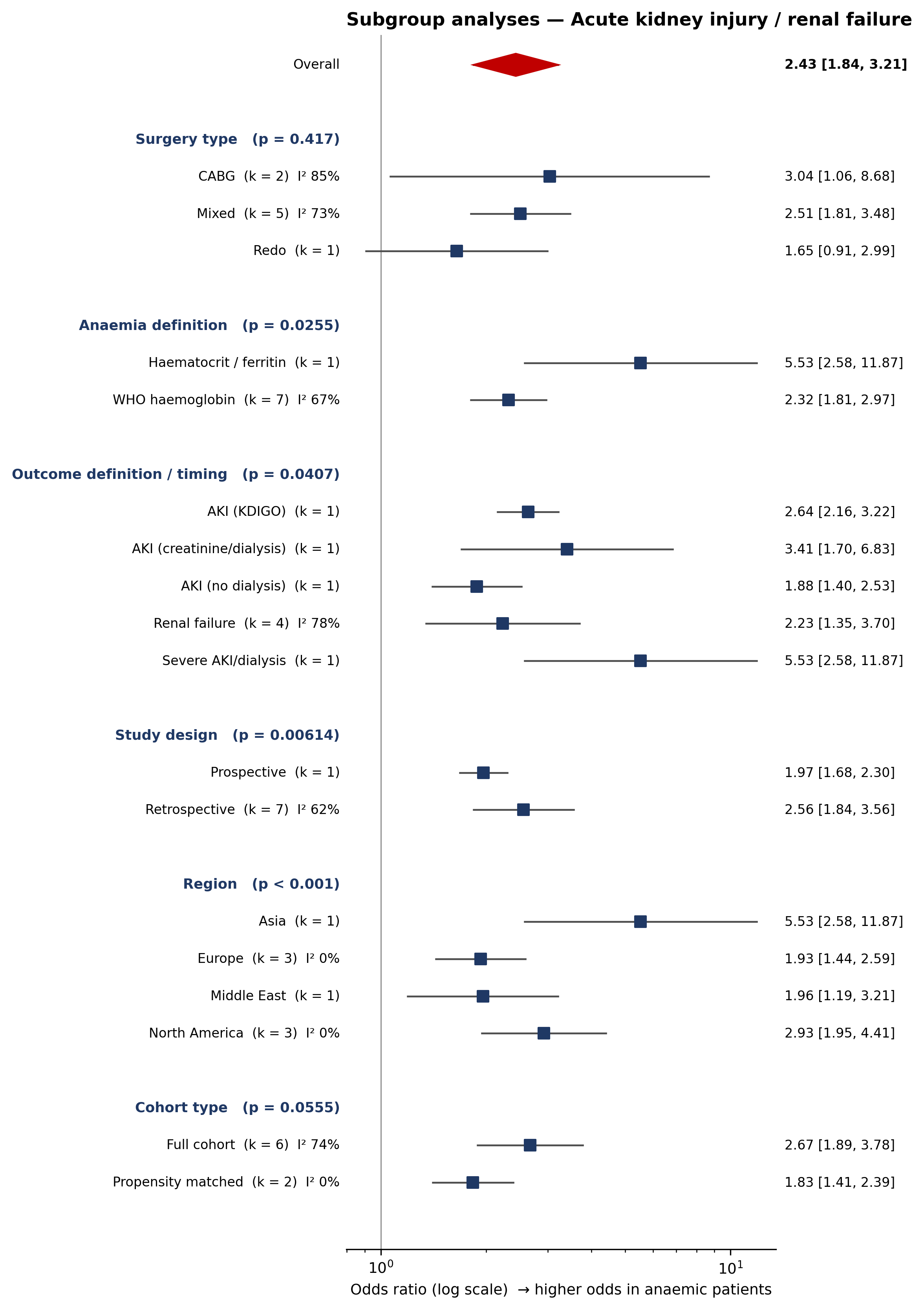

**Alt-text:** Forest plot of subgroup analyses for acute kidney injury or renal failure across eight studies. The overall odds ratio was 2.43 (95% CI 1.84–3.21; I² 70.1%), with subgroup analyses by surgery type, anaemia definition, outcome definition/timing, study design, region, and cohort type.

**eFigure S3: Subgroup analysis for early mortality**

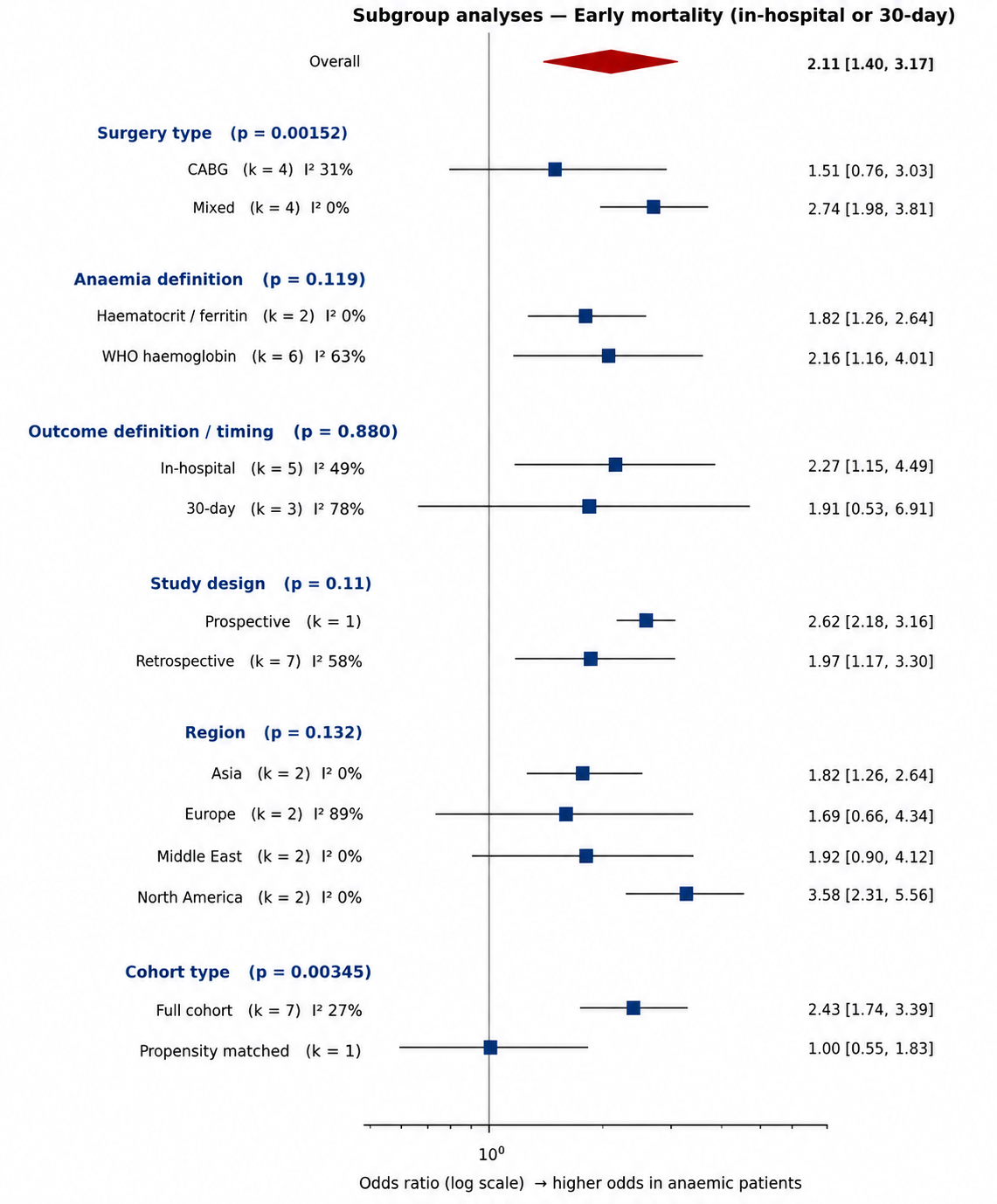

**Alt-text:** Forest plot of subgroup analyses for early mortality across eight studies. The overall odds ratio was 2.11 (95% CI 1.40–3.17; I² 58.4%), with subgroup analyses by surgery type, anaemia definition, outcome definition/timing, study design, region, and cohort type.

**eFigure S4: Subgroup analysis for postoperative infection**

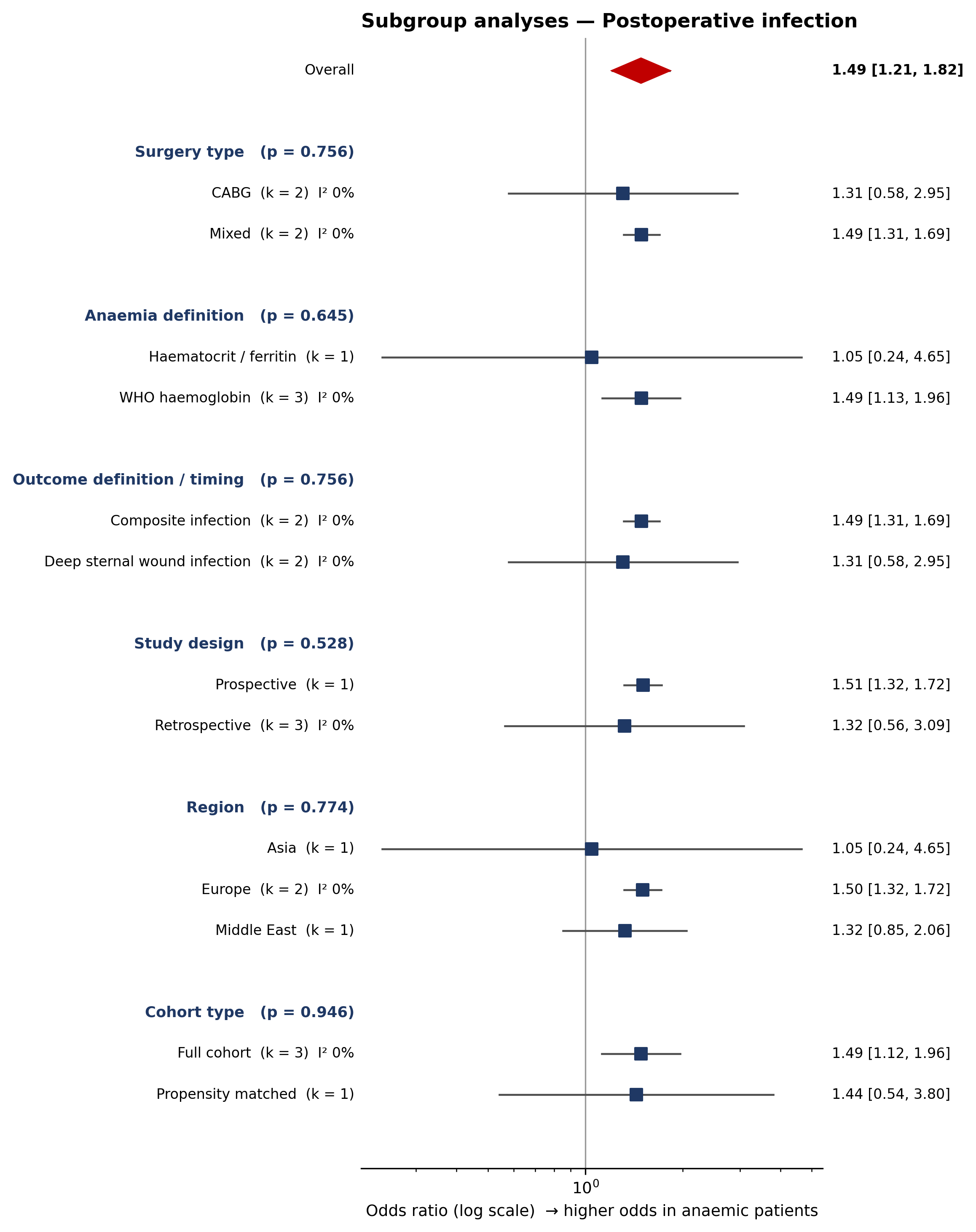

**Alt-text:** Forest plot of subgroup analyses for postoperative infection across four studies. The overall odds ratio was 1.49 (95% CI 1.21–1.82; I² 0%), with subgroup analyses by surgery type, anaemia definition, outcome definition/timing, study design, region, and cohort type.

**eFigure S5: Subgroup analysis for postoperative stroke**

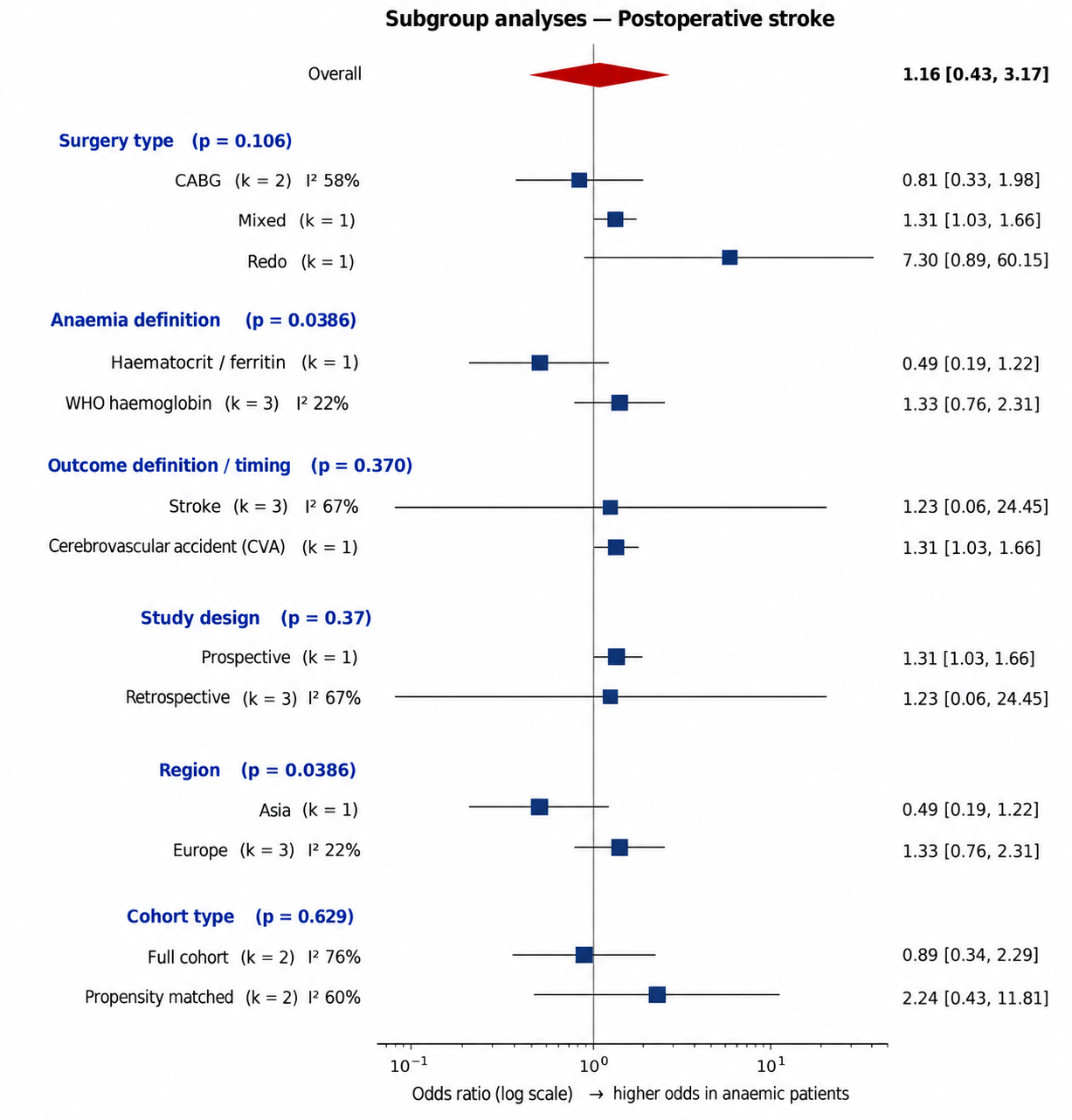

**Alt-text:** Forest plot of subgroup analyses for postoperative stroke across four studies. The overall odds ratio was 1.16 (95% CI 0.43–3.17; I² 56.3%), with subgroup analyses by surgery type, anaemia definition, outcome definition/timing, study design, region, and cohort type.

**eFigure S6. Baujat plots for heterogeneity contribution and influence**

**on the pooled estimates across the five postoperative outcomes**

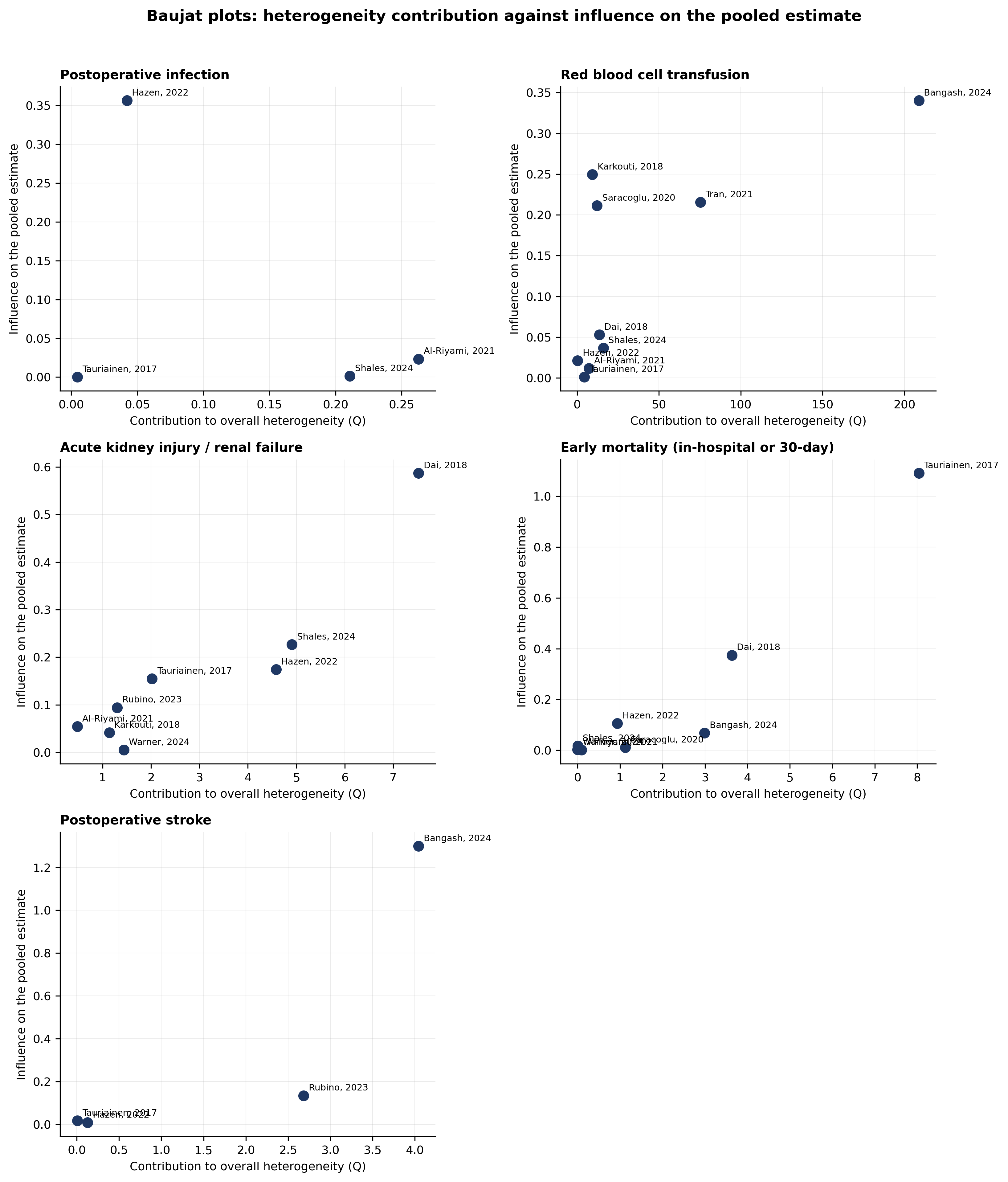

**Alt-text:** Baujat plots for postoperative infection, red blood cell transfusion, acute kidney injury or renal failure, early mortality, and postoperative stroke. Each point represents one study; the x-axis shows contribution to overall heterogeneity and the y-axis shows influence on the pooled estimate. Studies located farther toward the upper right contribute more to heterogeneity and have greater influence on the pooled effect.

**eFigure S7. Externally standardised residuals for the five postoperative outcomes.**

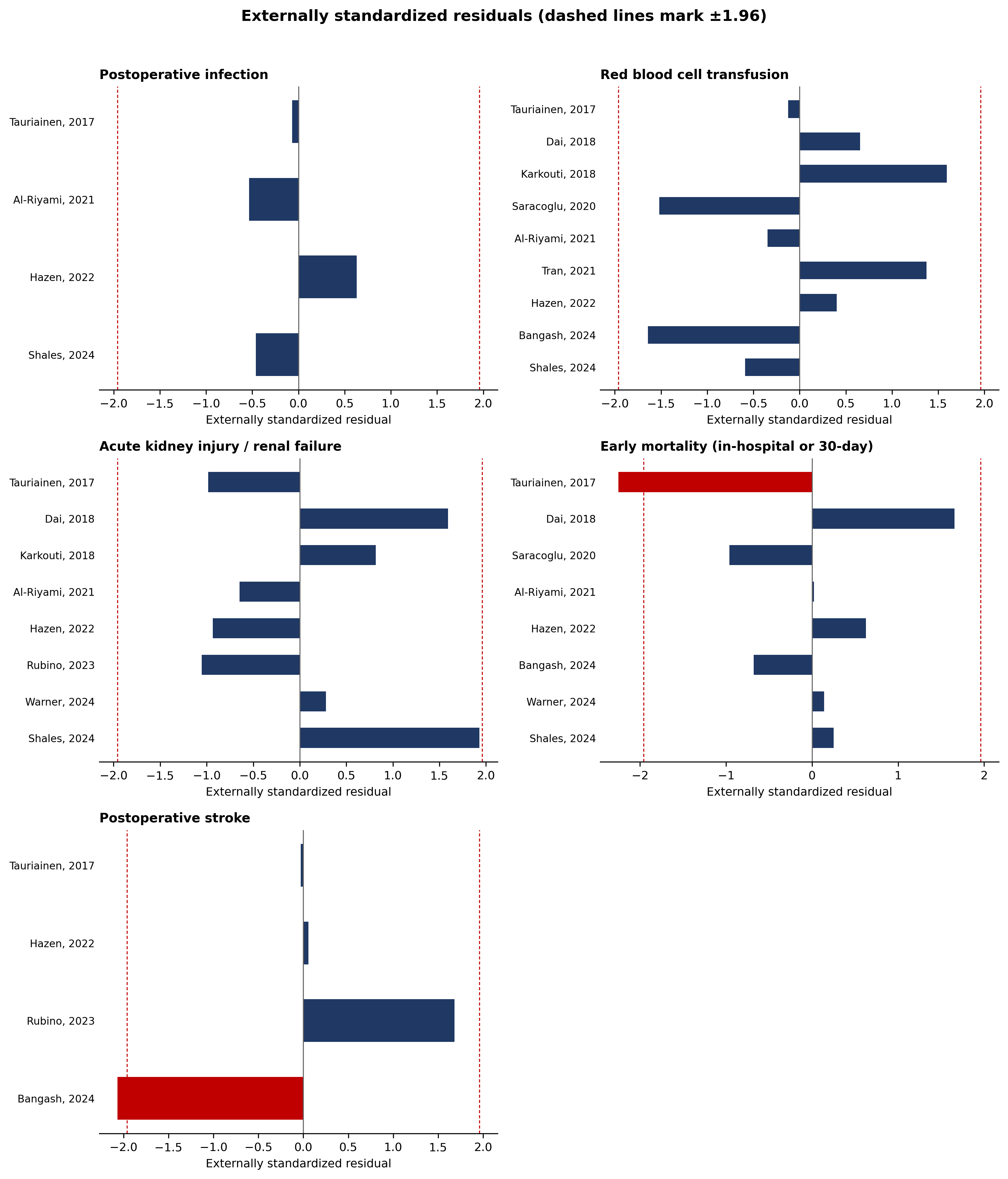

**Alt-text:** Externally standardised residual plots for the five postoperative outcomes. Each horizontal bar represents an individual study, and dashed vertical lines indicate thresholds of −1.96 and +1.96. Bars extending beyond these thresholds identify studies with potentially unusual residuals.

**eFigure S8. Baseline transfusion rate versus the observed odds ratio for red blood cell transfusion.**

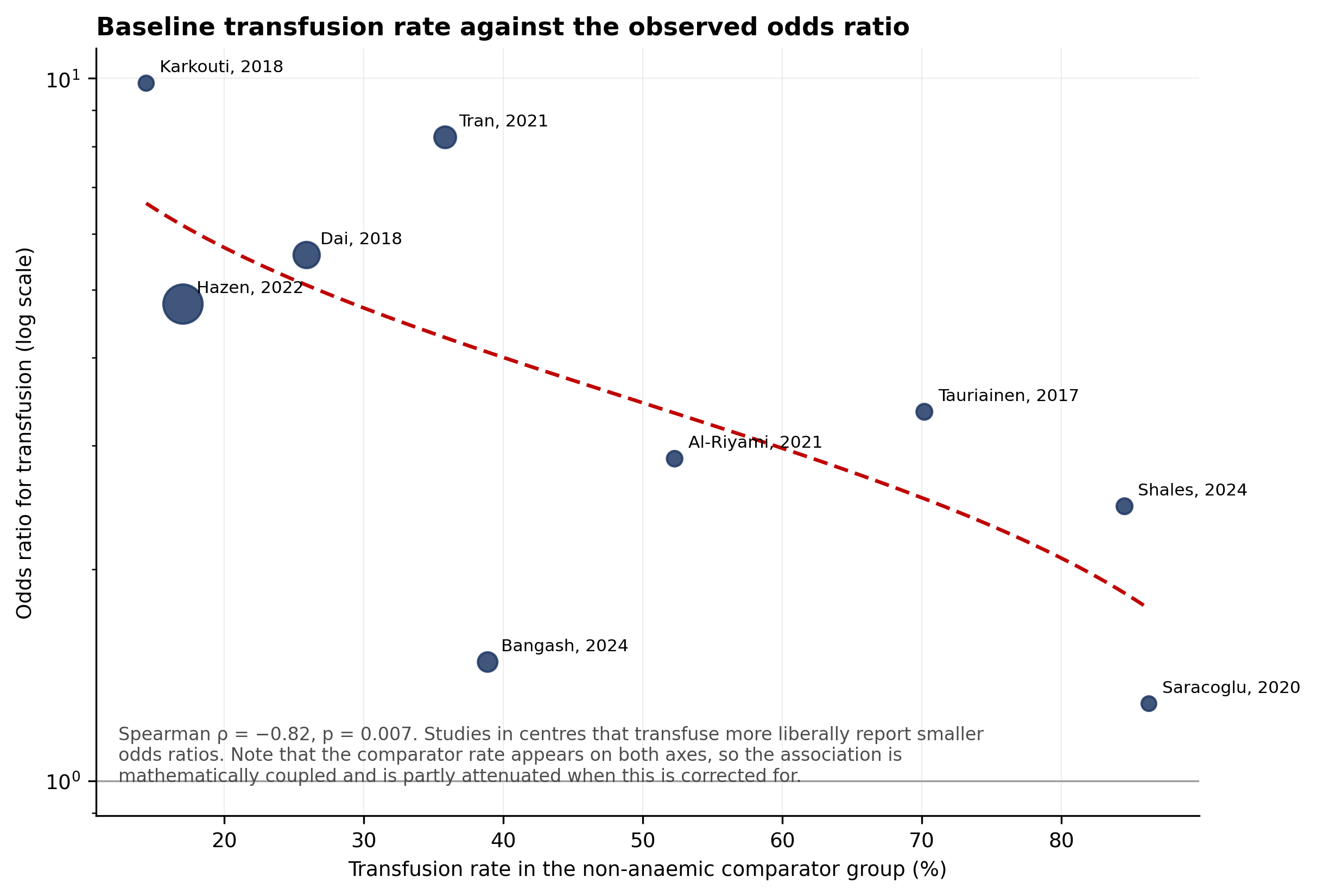

**Alt-text:** Scatter plot of nine studies showing the relationship between the transfusion rate in the non-anaemic comparator group and the observed odds ratio for red blood cell transfusion. Higher baseline transfusion rates were associated with smaller observed odds ratios (Spearman ρ = −0.82, p = 0.007). The dashed regression line shows the overall negative trend; the association should be interpreted cautiously because the comparator transfusion rate contributes mathematically to the odds ratio.
